# Hitting the diagnostic sweet spot: Point-of-care SARS-CoV-2 salivary antigen testing with an off-the-shelf glucometer

**DOI:** 10.1101/2020.09.24.20200394

**Authors:** Naveen K. Singh, Partha Ray, Aaron F. Carlin, Celestine Magallanes, Sydney C. Morgan, Louise C. Laurent, Eliah S. Aronoff-Spencer, Drew A. Hall

**Affiliations:** Department of Electrical and Computer Engineering, University of California – San Diego, La Jolla, CA 92093, USA; Division of Surgical Oncology, Department of Surgery, Moores Cancer Center, University of California – San Diego Health, La Jolla, CA 92093, USA; Division of Infectious Diseases and Global Public Health, Department of Medicine, University of California – San Diego, La Jolla, CA 92093, USA; Department of Obstetrics, Gynecology, and Reproductive Sciences, University of California – San Diego, La Jolla, CA 92093, USA; Department of Bioengineering, University of California – San Diego, La Jolla, CA 92093, USA

**Keywords:** aptamer, glucometer, SARS-CoV-2, CoVID-19, point-of-care, population screening

## Abstract

Significant barriers to the diagnosis of latent and acute SARS-CoV-2 infection continue to hamper population-based screening efforts required to contain the COVID-19 pandemic in the absence of effective antiviral therapeutics or vaccines. We report an aptamer-based SARS-CoV-2 salivary antigen assay employing only low-cost reagents ($3.20/test) and an off-the-shelf glucometer. The test was engineered around a glucometer as it is quantitative, easy to use, and the most prevalent piece of diagnostic equipment globally making the test highly scalable with an infrastructure that is already in place. Furthermore, many glucometers connect to smartphones providing an opportunity to integrate with contract tracing apps, medical providers, and electronic medical records. In clinical testing, the developed assay detected SARS-CoV-2 infection in patient saliva across a range of viral loads - as benchmarked by RT-qPCR - within one hour, with 100% sensitivity (positive percent agreement) and distinguished infected specimens from off-target antigens in uninfected controls with 100% specificity (negative percent agreement). We propose that this approach can provide an inexpensive, rapid, and accurate diagnostic for distributed screening of SARS-CoV-2 infection at scale.

## Introduction

Since the first reports of a deadly respiratory illness from Wuhan, China in the winter of 2019, Severe Acute Respiratory Syndrome Coronavirus 2 (SARS-CoV-2), the causative agent of COVID-19, has spread globally and resulted in the most impactful pandemic in more than a century. This pathogen joins a growing number of emerging infectious diseases, including Avian and H1N1 “swine” influenza, human immunodeficiency virus (HIV), Ebola viruses, Zika virus, Middle East respiratory syndrome Coronavirus (MERS-CoV), and severe acute respiratory syndrome (SARS) Coronavirus (SARS-CoV), which are increasing in frequency and severity as a consequence of human related activities such as globalization, societal unrest, and changes in environmental conditions^11^. To a greater or lesser extent, these viruses share important features, including passage through intermediate non-human hosts leading to genetic alterations that result in increased infectivity and virulence in human populations, asymptomatic carriage in a proportion of infected individuals allowing for extensive undetected spread, and inadequate availability of accurate diagnostic tests. Even as COVID-19 has reached over 214 countries, true prevalence remains difficult to estimate due to continued limitations in the capacity and performance of existing molecular diagnostics^2–4^. In this context, the number of confirmed cases exceeds 29 million and there have been over 920,000 attributed deaths to date. To date, the United States has the greatest number of cases of any country in the world (over 6.7 million as of 9/15/2020), or nearly 25% of global incidence^5^ and has conducted 92 million tests or ∼22% of the more than 2 billion viral tests that have been performed globally^6^.

There are currently three accepted methods for diagnosis of SARS-CoV-2 infection: 1) Viral RNA detection; 2) Viral protein detection, typically against the nucleocapsid (N) protein or spike (S) surface glycoprotein; and 3) Measurement of specific antibodies directed towards viral proteins. While the initial antibody response may be detected within a week of symptoms, (IgM as early as day 7 and IgG >14-days post infection)^7^, direct viral testing has been the preferred screening method in asymptomatic populations and acute presentations. For this, reverse transcriptase polymerase chain reaction (RT-PCR) remains the gold standard nucleic acid amplification test (NAAT), with samples collected from nasopharyngeal, mid-turbinate, oropharyngeal (saliva), and bronchoscopy specimens. While sensitivity and specificity in ideal settings approaches 99%, reported real-world sensitivities are estimated to be as low as 70%^8,9^, likely due to variation in sample collection methods and differences in viral shedding across the oropharynx and respiratory tract^10^. Importantly, it is now recognized that presence of RNA detected by PCR may not reflect infectivity^11^, potentially making viral antigen detection a more appropriate biomarker of transmissible disease^12^.

Tromberg and others have discussed the continued challenges in SARS-CoV-2 testing^13^. Current efforts are hampered by limited capacity, cost, and deployment logistics, leading to prioritized testing of specific high-risk groups and leaving many populations without the level of screening necessary to control the spread of infection^14^. Moreover, we still lack a “perfect test” with high sensitivity to rule in, high specificity to rule out, the ability to discern active and past infection, rapid turn-around-time, and a price-point to allow testing at scale. Ideally, such a test could be performed by an inexperienced user (*e*.*g*., at-home or in the community), be able to reliably detect early (asymptomatic or acute) infection with a low false positive rate, and have results that can be objectively read and easily transmitted to patients’ medical providers and public health personnel. Lateral flow immunoassays (LFIA) are a promising point-of-care (POC) solution, but are associated with important limitations, including qualitative readout, and reportedly low sensitivity with high false positive rates^15–17^. Hence, there remains an urgent need for accurate and cost-effective diagnostic tests that can be broadly deployed.

A potential solution to this problem is to develop quantitative, rapid tests around infrastructure that has already been adopted in the market at-scale. It has been previously shown that commercial glucometers can be re-purposed to detect a variety of non-glucose-based targets, quantitatively measuring cocaine, Ebola, hepatitis B, food borne pathogens, and interferon gamma^18–26^. There are currently 422 million people worldwide who rely on these devices daily to manage their blood sugar, making the glucometer the most prevalent piece of diagnostic equipment globally^27,28^. These meters are small, inexpensive ($20-50 USD), user friendly, highly accurate^29^, and many integrate with smartphones through Bluetooth, providing an opportunity to integrate SARS-CoV-2 detection results with contract tracing apps and medical providers. The major hurdle in repurposing a glucometer for direct detection of SARS-CoV-2 is that the target biomarkers (*e*.*g*., protein N and S) are present at low concentrations in biological samples The average CoVID-19 viral load in nasal/throat, sputum, and saliva samples is 3×10^6^, 7.50×10^5^, and 3.5×10^7^ copies/ml^30,31^, respectively, necessitating signal amplification to generate product (*i*.*e*. glucose) in quantities similar to physiological levels in human blood (*i*.*e*. 10–600 mg/dL or 0.6–33 mM)^27,32^.

Aiming to hit a SARS-CoV-2 diagnostic sweet-spot, we report a point-of-care saliva-based test that can quantitatively measure viral antigen with a glucometer. To transduce antigen binding events into glucose signal production, we exploit the native catalytic property of invertase and an aptamer-based competitive assay. The proposed workflow is illustrated in **Figure 1**, where aptamers directed at the viral S or N protein are pre-conjugated to invertase through a small antisense oligonucleotide strand that is complementary to a portion of the aptamer’s binding domain (aptatope). The biotinylated aptamer-oligo-invertase complex is pre-assembled on magnetic beads. In the presence of the conjugate antigen, the aptamer undergoes a conformational change, displacing the lower affinity antisense strand, thus creating an *antigen sensitive switch*. After magnetic separation, the released enzyme hydrolyzes sucrose into glucose with a turnover rate of 5×10^3^ glucose mol/sec^19^ enabling many orders of magnitude signal enhancement. This amplification allows readout with an off-the-shelf glucometer where the signal is proportional to the viral antigen concentration. In this work, we designed and optimized the system for saliva given the simplicity of sample collection^33^. However, this approach would work equally well with other sample types. Through testing, we demonstrate that the assay has minimal cross-reactivity to proteins from other respiratory viruses, recognizes native antigens in conditioned media of cells infected with SARS-CoV-2, and clinically discriminates infected and non-infected individuals with an unmodified $29 glucometer.

**Figure 1.**
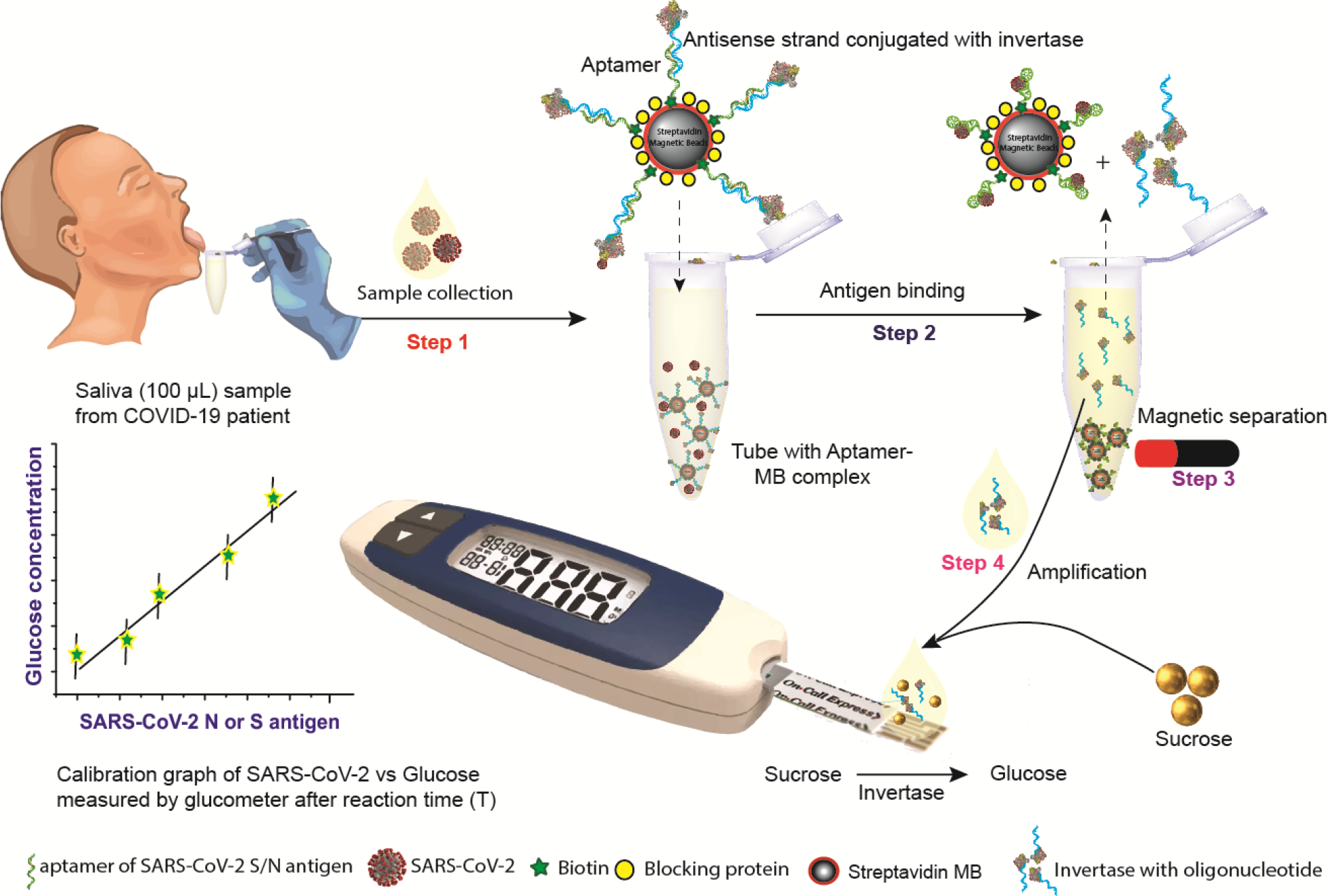
Overview of proposed point-of-care, aptamer-based COVID-19 assay. SARS-CoV-2 N or S protein specific biotinylated aptamer is conjugated to streptavidin coated magnetic bead (MB) and pre-hybridized with a complementary antisense oligonucleotide strand that is covalently attached to an invertase enzyme. The saliva sample is added to this cocktail (**Step 1**). Upon binding to the viral antigen or the SARS-CoV-2 virion, the invertase-antisense oligo is released (**Step 2**). A magnet is used to remove the MB-conjugated to the aptamer-antigen complex and the remaining aptamer-antisense-invertase complex (**Step 3**). The solution containing the released antisense-invertase is then collected and incubated with sucrose (**Step 4**). Invertase converts sucrose to glucose that is directly readout using a glucometer. The glucose concentration is correlated with the SARS-CoV-2 N or S protein concentration.

## Materials and Methods

### Reagents

Biotin-tagged, HPLC-grade purified aptamers against SARS-CoV-2 N^26^ and S^34^ antigen and complementary thiolated antisense DNA oligonucleotides were designed and ordered from Integrated DNA Technology (IDT). The aptamer and antisense sequences used in this work are listed in **Supplemental Table S1**. Streptavidin-coated Dynabeads M-280 (2.8 µm), 10% bovine serum albumin (BSA), dithiobis(succinimidyl propionate) (DSP), and tris(2-carboxyethyl) phosphine (TCEP) were purchased from Thermo Fisher. Dulbecco’s potassium phosphate buffer (DPBS) with calcium and magnesium, citrate buffer, calcium chloride (CaCl_2_), magnesium chloride (MgCl_2_), ethylenediaminetetraacetic acid (EDTA), sodium borohydride (NaBH_4_), sucrose, glucose, 4-(N-Maleimidomethyl)cyclohexane-1-carboxylic acid 3-sulfo-N-hydroxysuccinimide ester sodium salt (sulfo-SMCC), glucose oxidase type-VII from *Aspergillus niger*, and invertase (Grade VII) from *Saccharomyces cerevisiae* were purchased from Sigma Aldrich. All reagents were analytical grade and used without further processing. Buffer compositions are described in **Supplemental Table S2**. SARS-CoV-2 N and S, Influenza A (H1N1) hemagglutinin and neuraminidase, and MERS nucleocapsid and spike RBD fragment proteins were purchased from Sino Biological. Amicon filters (3, 10, and 100 kDa cutoffs) were purchased from Millipore. Dulbecco’s Modified Eagle’s Medium (DMEM) culture media and Penicillin-Streptomycin (10,000 U/ml) antibiotics were obtained from Corning and Gibco, respectively. An “Accu-Chek GuideMe” glucometer was used for all assays. The reagents with catalog numbers are listed in **Supplemental Table S3**.

### Conjugation of invertase with the antisense oligomer strand

Invertase was covalently linked with a thiolated antisense oligomer strand (specific to the N or S aptamer) using a modified version of a previously reported protocol^19^. Briefly, 30 µL of 1 mM thiolated antisense oligomer was mixed with 6 µL of 0.5 M TECP and stirred at room temperature (RT) for 2 hours. After incubation, the antisense strand was purified through centrifugation with a 3 kDa cutoff filter. This was repeated 8× in DPBS buffer. Next, 400 µL of invertase was mixed with 1 mg of water-soluble sulfo-SMCC by gentle pipetting for 5 min. The mixture was then placed on a shaker for 2 hours at RT. After incubation, unbound sulfo-SMCC was removed by centrifugation with a 10 kDa cutoff filter. This process was repeated 8× in DPBS buffer. The purified sulfo-SMCC linked invertase (sulfo-SMCC-invertase) was mixed with the purified, reduced, thiolated antisense strand and kept on a shaker for 48 hours at RT. Unreacted free antisense oligomers were removed by centrifugation with a 10 kDa cutoff filter. This was repeated 8× in DPBS. The purified antisense-invertase conjugate was stored at 4°C for downstream application.

### Hybridization of aptamer with antisense-invertase conjugate

The biotinylated (N or S) aptamer was refolded by heat treatment at 80°C for 3 min, followed by gentle cooling at RT for 5 min. Similarly, the antisense-invertase conjugate was heat treated at 40°C for 10 min before hybridization. 10 µL of heat-treated N or S aptamer (0.5 mM) was mixed with 20 µL of heat-treated antisense-invertase conjugate and 170 µL of DPBS and placed on a shaker for 2 hours at RT. The unhybridized free aptamer was removed by centrifugation with a 100 kDa cutoff filter. This was repeated 8× with washing in DPBS. The purified, hybridized aptamer/antisense-invertase complex (∼200 µL) was stored at 4°C.

### Conjugation of aptamer/antisense-invertase complex and magnetic beads

200 µL of streptavidin coated magnetic beads (MBs) was placed near a rare earth magnet. The supernatant was discarded and replaced with 600 µL of washing and binding buffer (see **Supplemental Table S2**). This process was repeated 3×. The MBs were then equilibrated with DPBS buffer for 10 min, the incubation buffer discarded, and resuspended in 200 µL of the biotinylated aptamer/antisense-invertase complex. This was kept on a shaker for 1 hour at RT. Excess unbound aptamer/antisense-invertase complex was washed off with buffer 3-5× times. The resulting aptamer/antisense-invertase magnetic bead complex (MBC) was treated with 1% BSA in DPBS for 30 min. After incubation, the BSA solution was discarded and the MBC was resuspended in 400 µL of DPBS. 50 µL of the MBC (∼200 µg) was aliquoted in test tubes and stored at 4°C.

### Fabrication of the custom electrochemical glucose sensor

A glass slide with an evaporated gold electrode (5 nm Ti / 50 nm Au) was chemically cleaned in piranha solution (3:1 of H_2_SO_4_:H_2_O_2_) for 1 min followed by washing with ultrapure (milli-Q) water. The electrode was then sonicated in acetone and isopropanol sequentially for 5 min followed by washing with ultrapure water. The electrode was then electrochemically cleaned in 0.5 M H_2_SO_4_ by sweeping the potential from −0.5 to +1.2 V vs. Ag/AgCl electrode, washed with water, and air dried. A surface assembled monolayer (SAM) was formed by incubating the electrode in 1 ml DSP (2 mg/ml) reduced with 5 µL of (10 mg/ml) NaBH_4_ for 2 hours at RT. The electrode was then washed with acetone, methanol, isopropanol, and ultrapure water followed by air drying. The DSP modified electrode was incubated with 5 µM of glucose oxidase in PBS overnight at 4°C to covalently link to the DSP modified surface. The unbound GOx was washed off with PBS and the sensor was incubated in 1% ethanol amine for 15 min to block any remaining active succinimidyl and then 1% BSA for 10 min. The electrode was stored at 4°C when not in use. Layer by layer assembly was monitored by cyclic voltammetry (CV) with a CHI-760E electrochemical workstation in a three-electrode configuration (BASi Ag/AgCl reference electrode and a platinum wire counter electrode). Voltammograms were measured from −0.5 to +0.8 V at a scan rate of 50 mV/s with 1 mM ferrocene in 1× PBS and 0.25 M KCl.

### Custom-made glucose sensor SARS-CoV-2 assay

100 µL of DPBS buffer spiked with SARS-CoV-2 N or S protein was incubated with 200 μg of MBC (N or S) with gentle shaking for 30 min at RT in a 1.5 ml centrifuge tube. The MBC was pulled down using a rare earth magnet. 90 μL of the supernatant was transferred to another centrifuge tube prefilled with 100 μL of 2× Measurement buffer (**Supplemental Table S2**). After incubating for 30 min, 200 µL was placed on the glucose sensor and readout using a CHI-760E electrochemical workstation with the three-electrode configuration described previously. Voltammograms were measured from −0.5 to +0.8 V at a scan rate of 50 mV/s.

### Collection of nasopharyngeal swab and saliva samples

We collected matched nasopharyngeal swabs (NPS) in RNAShield DNA/RNA storage medium (Zymo) or viral transport medium and saliva samples (no additive) from symptomatic and asymptomatic study subjects with a prior positive clinical COVID-19 RT-qPCR result under IRB approval (UCSD protocol #200477). These samples were subjected to viral RNA extraction using the MagMax Viral/Pathogen Nucleic Acid Isolation Kit (Thermo) and the TaqPath COVID-19 multiplex RT-qPCR assay was performed on the resulting RNA samples. Saliva in 300 µL aliquots were provided as blinded specimens for testing in the BSL3 lab. Demographic information about the cohort is listed in **Supplemental Table S4**.

### Glucometer-based SARS-CoV-2 assay

100 µL of sample (contrived, conditioned media, or saliva) was diluted two-fold in DPBS buffer. Half of the diluted sample was then incubated with 200 μg of MBC (N or S) with gentle shaking for 30 min at RT in a 1.5 ml centrifuge tube. The MBC was pulled down using a rare earth magnet. 90 μL of supernatant was transferred to a centrifuge tube prefilled with the Sucrose buffer (**Supplemental Table S2**). The other half of the diluted sample was placed in a separate centrifuge tube prefilled with the Sucrose buffer as a background control. After mixing, the tubes were incubated at 60°C in a water bath for 1 hour. Finally, 10 μL of each reaction solution was placed on a glucometer test strip and read out using a glucometer. The difference between the two readings was recorded. All measurements were repeated in triplicates.

### Authentic SARS-CoV-2 production and quantification

Vero E6 cells were obtained from ATCC and grown in DMEM with 10% fetal bovine serum (FBS) and Penicillin-Streptomycin. SARS-CoV-2 isolate USA-WA1/2020 (BEI Resources) was propagated and aliquots of secreted virus in culture media were stored at −80°C. Infectious units (IU) were quantified by digital droplet PCR (ddPCR) and plaque assay using Vero E6 cells. For ddPCR, viral stock media was added to TRIzol LS (ThermoFisher) and RNA extracted using a Directzol RNA miniprep kit (Zymo Research). The ddPCR quantified SARS-CoV-2 *ORF1a* and was performed by the UCSD Center for Aids Research (CFAR) Genomics and Sequencing Core. For plaque assay quantification, viral supernatants were 10-fold serially diluted in DMEM without serum. Vero E6 cells in 12-well plates were washed with PBS, and 200 μL of virus dilution was added per well and incubated 1 hour at 37°C with rocking every 10-15 min. The inoculum was removed and 1 ml of overlay (0.6% agarose in MEM with 4% FBS) was added to each well. Overlays were prepared by mixing equal volumes of 1.2% agarose and 2× MEM supplemented with 8% FBS, 2× L-glutamine, 2× non-essential amino acids, and 2× sodium bicarbonate. Assays were incubated for 48 hours at 37°C and fixed by adding 2 ml 10% formaldehyde for at least 24 hours. Overlays were removed, and monolayers were stained with 0.025% crystal violet in 2% EtOH and plaques counted.

### Safety

Piranha solution is highly corrosive and extreme precaution is needed in the handling. All work involving infectious SARS-CoV-2 samples was undertaken in the UC San Diego Division of Infectious Diseases Biosafety Level 3 (BSL3) laboratory with oversight from the UC San Diego Institutional Biosafety Committee (IBC).

### Statistical analysis

All data was from a minimum of three independent experiments. Error bars represent one standard deviation. Statistical analysis was performed with Origin 9.0 and/or MATLAB. The limit of detection (LOD) was calculated using the slope method where LOD = 3×standard deviation (SD) of blank/slope^35^.

## Results

### Antigen-sensitive aptamer switch validation

We selected aptamers reported in the literature to have high affinity towards SARS-CoV-2 N and S antigen. We then analyzed their secondary structures using Mfold^36^ and designed corresponding antisense oligonucleotide strands to overlap the predicted secondary stem-loop structures. The 5′ end of the aptamers and antisense strands were extended with a linker (6 and 12 thymine oligomers, respectively) to increase the distance between the aptamer and the magnetic beads, allowing the aptamer room to properly fold and reduce steric hindrance. To test the release of the antisense strand from the aptamer upon ligand binding, we designed a PCR-based assay where protein binding induces a conformation change in the aptamer, releasing the antisense oligo, as shown in **Supplemental Figure S1**. The free antisense oligo was then collected from the supernatant and used as the reverse primer in a PCR reaction with the S and N aptamer as the template and the corresponding forward primers (**Supplemental Table S5)**. We confirmed PCR amplification through agarose gel electrophoresis. These data demonstrated specific antigen-mediated release of the antisense oligonucleotide from the aptamer and established baseline conditions for the subsequent assay development.

### Tuning assay conditions for glucometer read out

We carried out conjugation of the antisense oligo to invertase via amine functionalization to preserve the aspartic and glutamic acids in the enzyme’s active site^37^. Crosslinking the antisense oligonucleotide with invertase was performed with sulfo-SMCC and evaluated by an electrophoretic mobility shift assay (EMSA). The results, shown in **Supplemental Figure S2**, indicate successful crosslinking between the DNA and invertase, as visible from the gel image. We verified enzyme activity in the presence of the substrate (sucrose) with a redox mediator and quantified the amount of glucose using custom-made glucose sensors that have higher sensitivity and dynamic range than glucometers (**Supplemental Figure S3**). As enzyme activity is dependent on various factors, we assessed different incubation temperature, substrate concentrations, buffers, pH, and salts to optimize for efficiency and linearity (**Supplemental Figures S4 and S5)**. Based on our findings, the optimum conditions for the amplification phase are 1 M sucrose, 5 mM CaCl_2_, 1 mM MgCl_2_, and 0.5 mM EDTA in 0.1 M citrate buffer at pH 5.0. Maximum invertase activity was observed at 60°C, roughly 6× higher than at room temperature. We then conjugated the aptamer to magnetic beads using streptavidin-biotin chemistry and optimized the magnetic bead to aptamer ratio to avoid overcrowding, which causes steric hindrance reducing antigen binding efficiency. We found that magnetic beads saturated with a ratio of 1:5 for both the N and S directed aptamers (**Supplemental Figure S6**). We performed studies to identify the optimum aptamer-target interaction binding time and signal amplification (invertase) time (**Supplemental Figure S7**). We selected 30 min for both. These optimized conditions were used for all subsequent assays.

### Quantitative SARS-CoV-2 protein detection in buffer and saliva

We measured the correlation between glucose readout and antigen concentration by spiking SARS-CoV-2 N and S antigen in buffer and healthy saliva (**Figure 2**). A calibration plot was generated by varying the antigen concentration across the same range as commercially available SARS-CoV-2 ELISA kits^38,39^. Using off-the-shelf glucose test strips and a glucometer, we observed a broad linear dynamic range of 1-500 pM for all combinations of sample matrix and antigen. The LODs in buffer were 1.50 pM and 1.31 pM for protein N and S, respectively. In saliva, the LODs increased to 4.38 pM and 5.76 pM for protein N and S, respectively. We further evaluated the assay performance with custom-made glucose sensors (**Supplemental Figure S8**). The custom electrochemical sensor performed similarly but achieved lower LODs (0.71 pM and 0.34 pM for protein N and S, respectively) due to the use of a high performance, benchtop potentiostat. Despite using a glucometer, this assay showed similar performance to ELISA kits for both N (3.5-226 pM) and S (2-128 pM) proteins.

**Figure 2.**
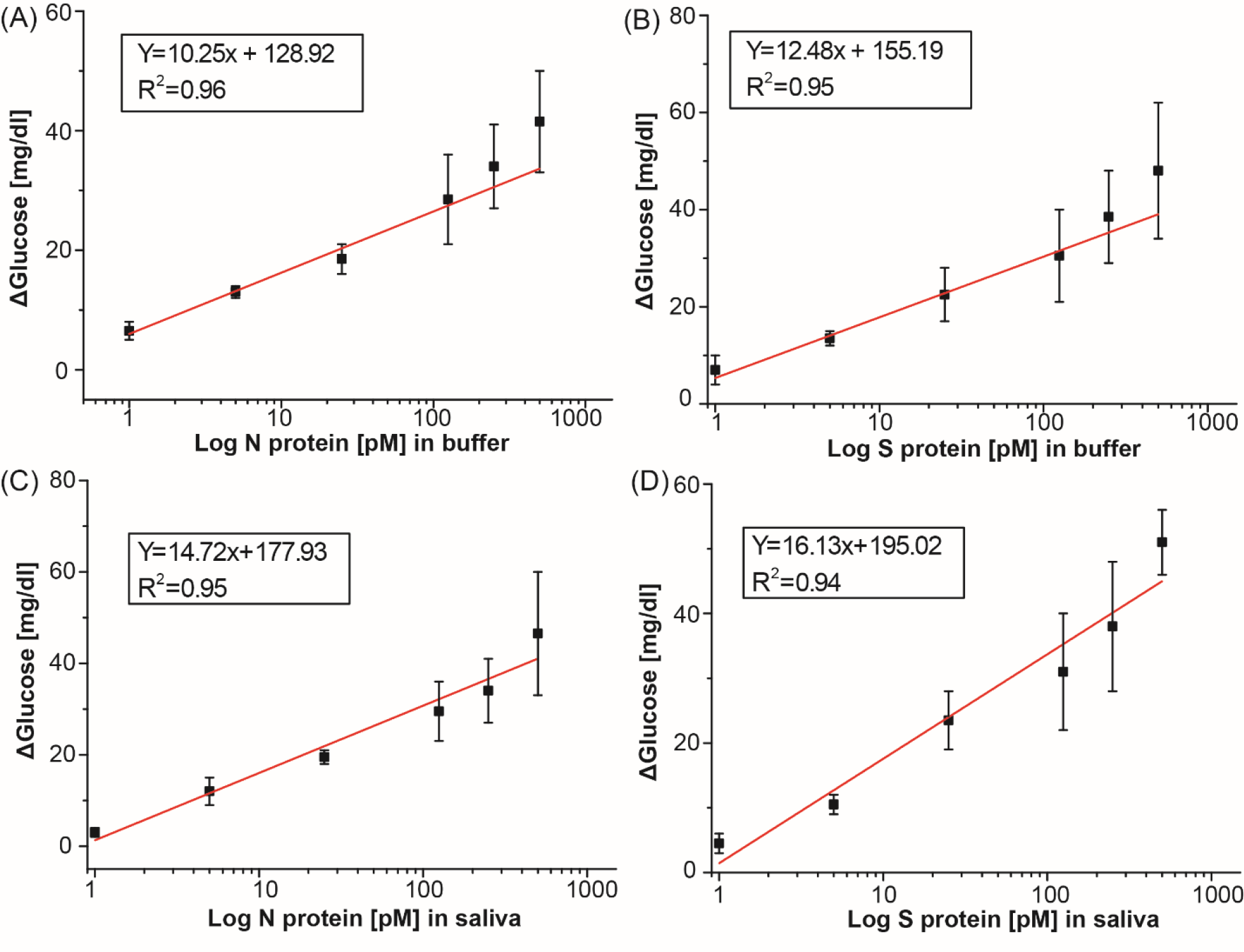
Calibration curves for SARS-CoV-2 N and S protein in buffer and saliva measured with a glucometer. Linear calibration curve with subtracted background signal of protein N spiked into (A) buffer and (C) saliva and protein S spiked in (B) buffer and (D) saliva.

### Determination of assay specificity

As a preliminary assessment of assay specificity, we measured signal generation in the presence of antigen from non-SARS-CoV-2 respiratory viruses, namely: Influenza A (H1N1) and MERS-CoV. Protein N and S specific aptamer complexes were assayed with the off-target antigens at a fixed concentration of 500 pM with the conditions described above. This assay was performed in buffer rather than saliva to isolate the source of the non-specific binding. The results, shown in **Figure 3**, indicate minimal cross-reactivity in the assay, even when the off-target antigens are present in high concentration. Unsurprisingly, the SARS-CoV-2 N aptamer displayed the highest signal with the MERS nucleocapsid antigen, and the S aptamer with the MERS-CoV RBD antigen, consistent with reported homology between the two coronavirus genomes. However, in all cases, the corresponding SARS-CoV-2 signal is >300% higher than the off-target recording (p < 0.05).

**Figure 3.**
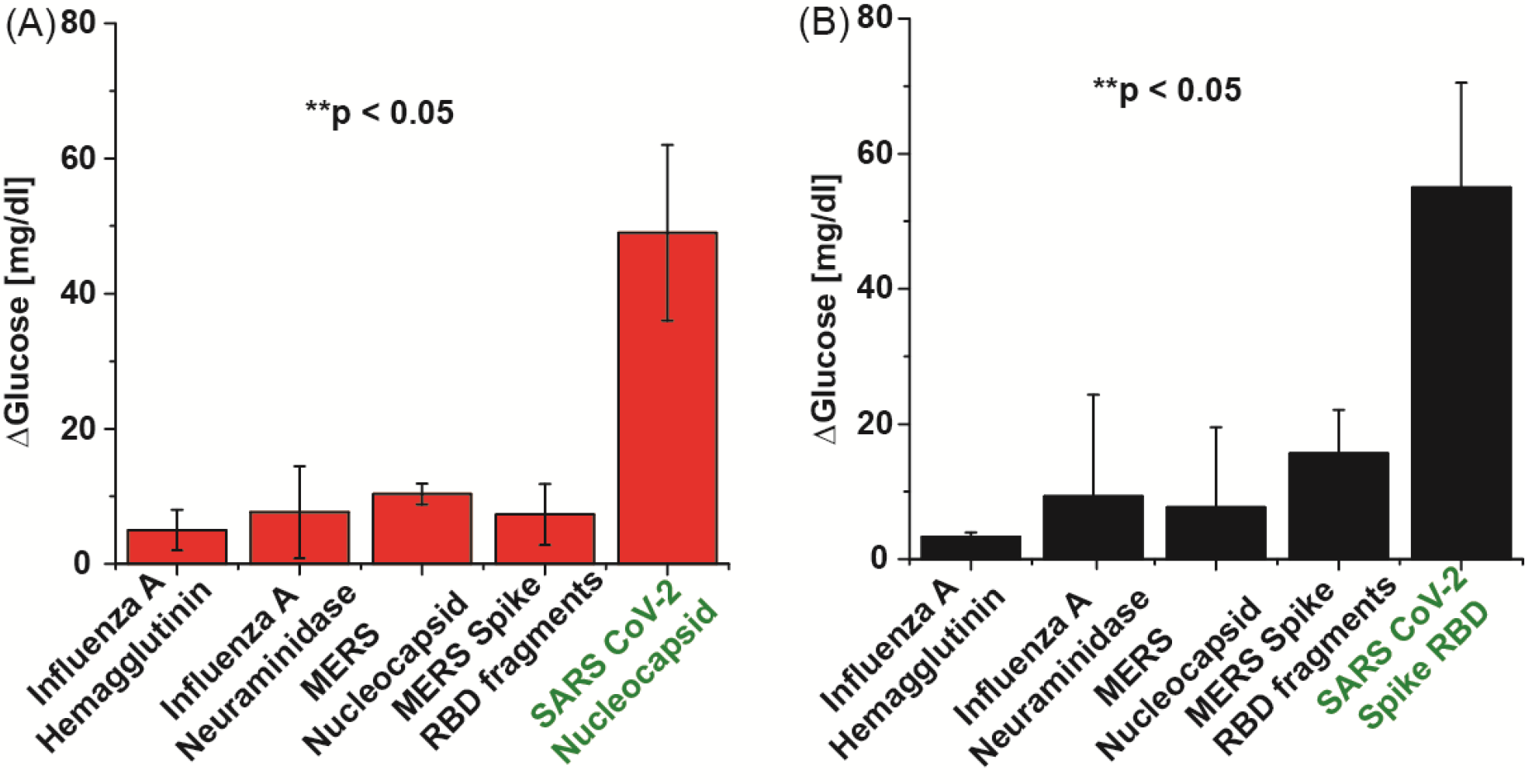
Cross-reactivity study. Assays using analogous proteins with (A) N aptamer complex and (B) S aptamer complex. All proteins were spiked into DPBS at 500 pM.

### Detection of authentic SARS-CoV-2 N and S protein in cultured media

Previous studies of protein S and N directed aptamers validated binding with only recombinant purified proteins^26,34^. To determine if our S and N aptamer/antisense-invertase system could recognize authentic virus and native proteins produced during SARS-CoV-2 infection, we created and quantified viral stocks of SARS-CoV-2 in a biosafety level 3 (BSL-3) laboratory. We inoculated authentic SARS-CoV-2 isolate USA-WA1/2020 onto Vero E6 cells and allowed these to propagate and secrete virus into the supernatant (**Figure 4A**). Supernatant was collected, aliquoted, and frozen. The quantity of SARS-CoV-2 in these preparations was determined by two methods: 1) we measured the quantity of SARS-CoV-2 O*RF1a* RNA using ddPCR, and 2) we determined the number of infectious virions by plaque assay. SARS-CoV-2 supernatants were diluted 1:10 with DPBS to 52×10^6^ copies and 12.5×10^3^ IU and assayed with the S and N aptamer complex. The cell media used to propagate the virus has a high level of glucose (450 mg/dl). To nullify the effect of background glucose in the conditioned media, we also conducted glucometer readings with control diluted media in a similar manner, but in the absence of the aptamer complexes. Both the N and S aptamers showed significant increases in glucometer readings compared to control samples (**Figure 4B**). This demonstrates that the aptamer/antisense-invertase systems recognize their native targets when produced by replicating authentic SARS-CoV-2.

**Figure 4.**
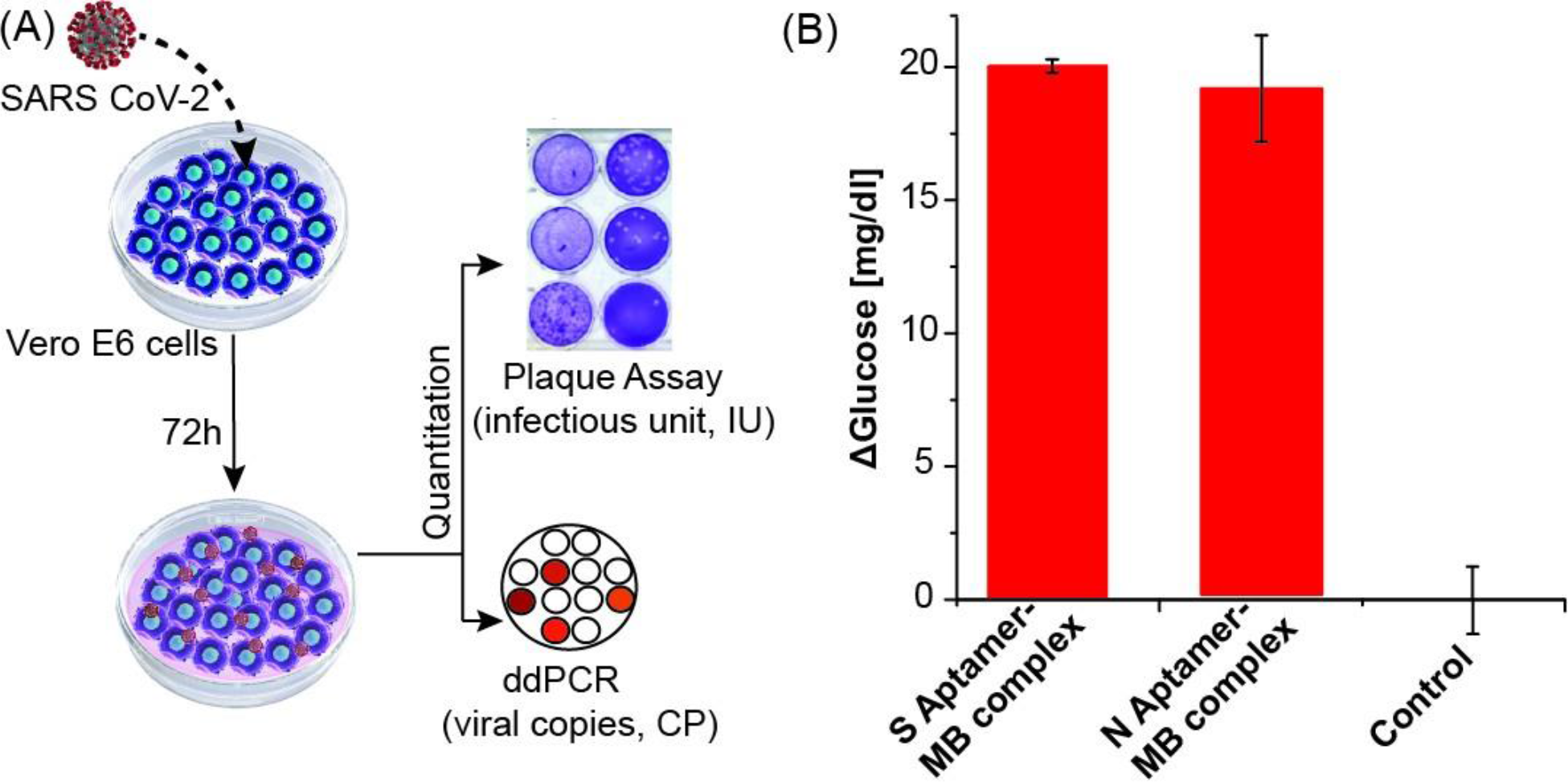
Detection of protein N and S using authentic SARS-CoV-2. (A) Schematic of authentic SARS-CoV-2 virus preparation and quantification of viral RNA by ddPCR and infectious units by plaque assay. (B) N and S aptamer/antisense MB complex detection of the SARS-CoV-2 native protein in 1:10 diluted virus culture media. Background media (control) values were subtracted from the measurement results.

### Detection of SARS CoV-2 in clinical specimens

Finally, we evaluated whether the developed assays could discriminate between SARS-CoV-2 infected and healthy individuals with validated saliva samples (see Materials and Methods). We started with a small cohort of 3 infected persons (confirmed positive with RT-qPCR) and 4 healthy controls tested for protein S and N binding. The results, shown in **Supplemental Figure S9**, demonstrate the ability of both assays to correctly differentiate between infected and non-infected individuals; however, the protein S assay showed significantly higher signal-to-control than the protein N assay and was selected as the focus of the larger study. In this cohort of 24 individuals, 42% were female and the average age was 31 years. Of the 16 infected individuals, the average time between symptom onset and testing was 7 days, 63% had a fever, and 50% had cough. Two subjects self-reported having asthma, one was pregnant, and none were diabetic. **Figure 5** shows the results that were presented as a blind panel run under BSL3 conditions over the course of two days using the same glucometer and a single lot of commercial test strips. All SARS-CoV-2 confirmed positive samples demonstrated higher glucose production (μ = 218 mg/dl, range = 68-404 mg/dl) than healthy individuals (μ = 24 mg/dl, range = 14-37 mg/dl). Receiver operator curve (ROC) analysis yielded an ideal cutoff of 52 mg/dl, which classified positive and negative samples with a sensitivity and specificity of 100% (AUC = 0.9988), as shown in **Supplemental Figure S10**. These data have 100% positive percent agreement (PPA) and 100% negative percent agreement (NPA) with the RT-qPCR data performed on the same samples (**Supplemental Table S5**).

**Figure 5.**
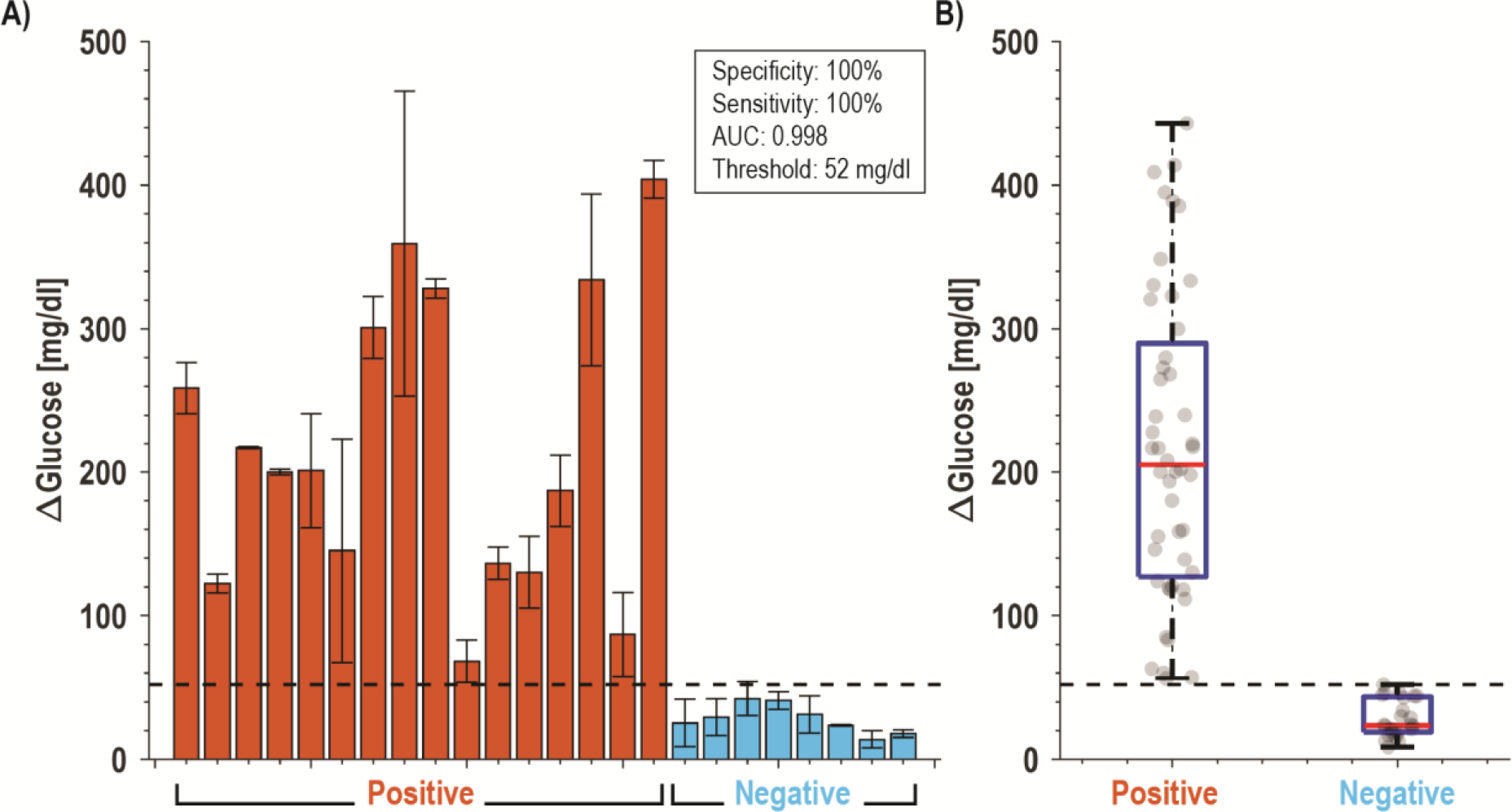
Clinical performance of saliva samples from CoVID-19 patients (*N*=16) and healthy volunteers (*N*=8). (A) Bar plot of all subjects using S aptamer magnetic bead complex and (B) box and whisker plot of all data points. With a cut-off threshold of 52 mg/dl, this assay has 100% PPA and 100% NPA with RT-qPCR data.

## Discussion

Based on the need for rapid, accurate, and easily scalable tests that can detect acute SARS-CoV-2 infection in large populations^40^, we developed and validated a novel aptamer-based sensor (an aptasensor) capable of sensitive detection of virus in human saliva using only low-cost reagents and “detectors” that are inexpensive and already ubiquitous worldwide – glucometers. The assay design required integration and optimization of three novel components: to bind antigen in human samples, to transduce binding into signal, and to detect that signal. There were several considerations that determined the selection of an aptamer affinity reagent over more commonly used protein-based molecules such as classical, single-chain, or camelid antibodies^41^. Aptamers are oligonucleotide (DNA or RNA) ligands that are selected through an iterative process known as SELEX^42,43^. They share similar affinity and specificity to monoclonal antibodies, yet can be mass produced at low cost, are stable at ambient temperatures for long-term storage, and can be chemically modified and engineered to produce conformational switches^44–46^. Specifically, the amount of overlap between the aptatope and the antisense strand in this assay allows the affinity to be engineered where it is intentionally designed to have low affinity for facile release when the viral protein is present. Notably, the sequence-based design of antigen dependent aptamer switches cannot be easily recapitulated with antibodies.

To develop a test that could be fielded for clinical use in months rather than years, we selected aptamers in the literature that had been previously validated for SARS-CoV-2 affinity. This allowed us to employ the second key element, a signal transduction and amplification step that converts viral antigen binding to signal production. Here the aptamer switch is coupled to invertase, a high efficiency sucrose converting enzyme which produces signal, glucose, in physiologic ranges. In this assay we chose an invertase from *S. cerevisiae* that has a high turnover rate to amplify biomarkers from low concentrations to those that could then be read by a glucometer. However, this enzyme has a peak efficiency near 60°C, which requires a heating element. While other POC tests have also used higher temperature to increase the assay kinetics^47,48^, there is potential to remove the heating element and to shorten time-to-answer with a room-temperature invertase from *Bacillus sp*.^49^ or other glucose converting enzymes.

The choice of saliva as a target specimen for testing presents challenges and opportunities. Saliva is the most readily obtained human specimen; however, it is also a complex and viscous sample consisting of various electrolytes, enzymes, and antibodies^38,39^ posing hurdles for specific and interference-free biomarker detection^40^. This is reflected in our data, where we found a higher LOD for protein spiked in saliva as compared to buffer, likely a consequence of the viscosity and proteases found in saliva. While invertase is not present in saliva, glucose is, especially after eating and/or drinking, which would interfere with the proposed readout method. Even with fasting, there will be some background glucose in saliva which could confound the measurement. To mitigate this, we dilute the saliva sample two-fold to reduce the effect from possible interferents and take a differential measurement to account for the initial (background) glucose and “signal leakage” from the low affinity antisense strand. This differential sampling would also account for persons with diabetic ketoacidosis (DKA). If using nasal swabs (nasopharyngeal or mid-turbinate), differential sampling would likely not be necessary.

Next, we tested the ability of the assay to detect native viral antigens during authentic cellular infection. Here we found significant differences between measurements of infected and uninfected viral media. These tests gave assurance that findings determined using free antigen in buffer and saliva could be replicated with native antigens, setting the stage for clinical testing. We then tested a blinded clinical panel comprising 24 saliva samples in a BSL3 facility. With a cutoff of 52 mg/dl determined by ROC analysis, all positive samples were discriminated from negative samples, giving a 100% level of agreement (LOA) with PCR testing and sensitivities and specificities of 100%. For each clinical sample, we correlated the mean glucose signal to patient and sample characteristics. Contrary to prior reports referencing better performance of antigen tests in specimens with high viral loads, we found no correlation (R <0.2) between the Ct value obtained from PCR and the reported test, nor did we find any relationship between days since onset and testing or clinical symptoms.

While at first glance, this would seem to indicate no quantitative relationship between infectious viral load and signal level, there are key pieces of missing data. First, PCR detects circulating nucleic acid which may be a poor proxy for infectiousness. Second, there is evidence of wide variation between nucleic acid copy number and viral antigen levels^50,51^, which may complicate comparison of detected antigen and nucleic acid levels. Likewise, in this study we chose to explore protein S, rather than the more commonly reported nucleocapsid protein. Each of these may have different measured correlations to viral load, and depending on site and state of infection, each may be accessible at different levels due to the presence of blocking antibodies. Even as it misses some resolved infections, salivary antigen detection has thus been suggested as a more appropriate for population testing than NAATs as it may be more likely to diagnose truly infectious individuals^33,52^. A definitive study, which is beyond the scope of this work, would be to use quantitative methods to measure antigen concentration in the samples, and further, measure intact viral genomes concomitantly with *in vitro* models to measure intact, infectious virions.

Since our test consists of readily available reagents and low-cost glucose test strips, each test has a modest $3.20 USD (see **Supplemental Table 2**) production cost with the current low volume production. It is worth noting that nearly 70% of the cost is the magnetic beads, which could be replaced with polystyrene beads and separated with a size selection filter, possibly in a syringe, rather than magnetic separation to reduce cost. Assuming that it is viable to reduce this to below $1 USD, our approach could provide a price point needed for population screening and repeated testing in both well- and under-resourced settings. Moreover, the numeric readout can be transmitted electronically allowing test reporting and tracking. With minor workflow improvements, a test such as this could be conducted at home, in dormatories, nursing homes or other ambulatory settings with support for remotely-observed-testing, digital reporting, and results notification via telemedicine or a smartphone app.

To situate our assay in the landscape of available SARS-CoV-2 diagnostics, **Table 1** shows a summary of reported point-of-care diagnostic tests. Here we see a diversity of approaches across the spectrum of nucleic acid and antigen modalities. However, there are notable gaps, including a paucity of saliva-based antigen tests and a complete lack of tests that provide a quantitative readout. The reported test fills this gap without sacrificing the sensitivity or specificity, using devices that already exist at scale with an easily acquired sample.

**Table 1.**
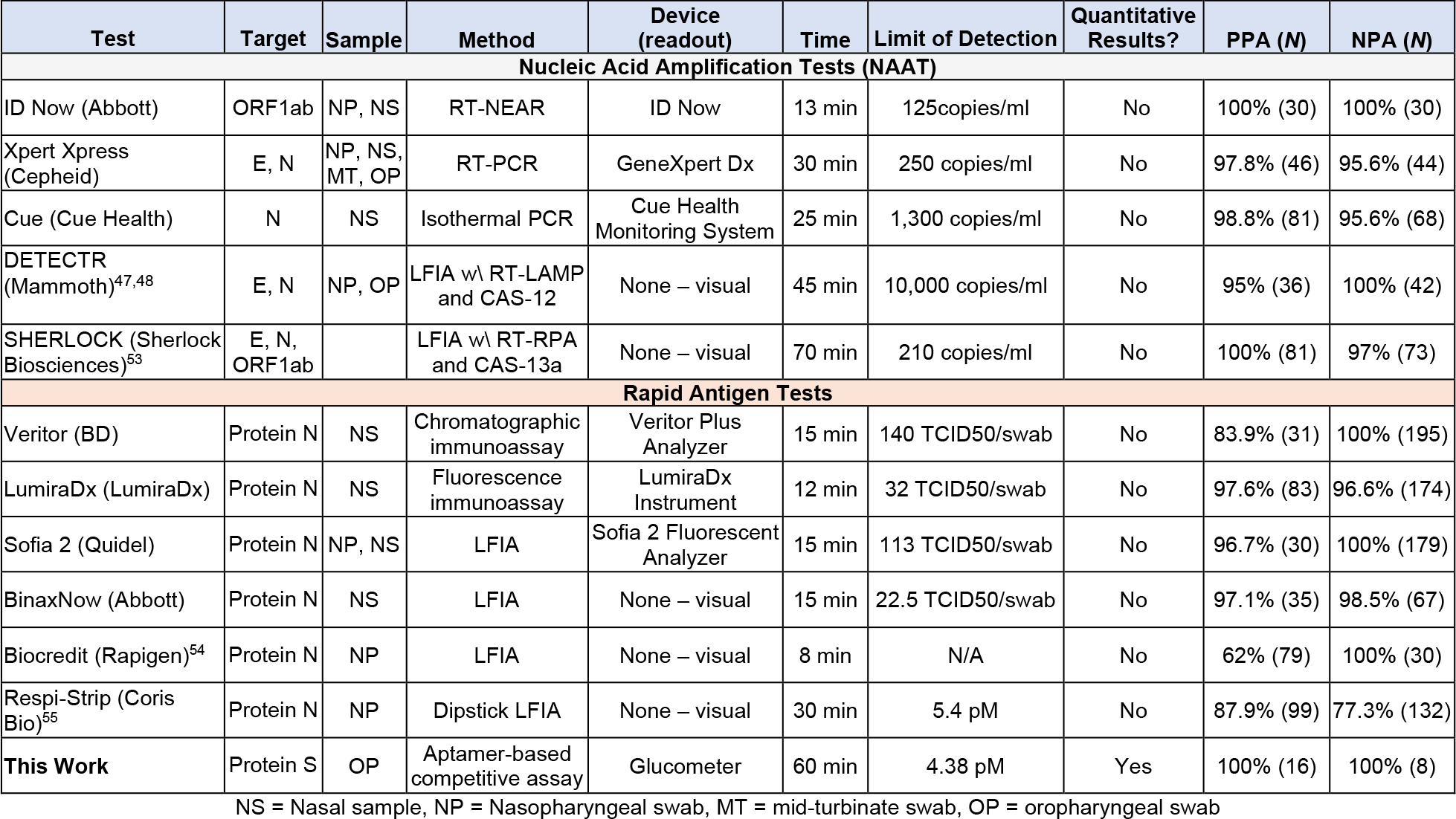
Comparison of Point-of-Care SARS-CoV-2 diagnostic tests. Unless otherwise noted, data compiled from the FDA Emergency Use Authorization (EUA) submission.

| Test | Target | Sample | Method | Device (readout) | Time | Limit of Detection | Quantitative Results? | PPA (N) | NPA (N) |
| --- | --- | --- | --- | --- | --- | --- | --- | --- | --- |
| **Nucleic Acid Amplification Tests (NAAT)** |  |  |  |  |  |  |  |  |  |
| ID Now (Abbott) | ORF1ab | NP, NS | RT-NEAR | ID Now | 13 min | 125copies/ml | No | 100% (30) | 100% (30) |
| Xpert Xpress (Cepheid) | E, N | NP, NS, MT, OP | RT-PCR | GeneXpert Dx | 30 min | 250 copies/ml | No | 97.8% (46) | 95.6% (44) |
| Cue (Cue Health) | N | NS | Isothermal PCR | Cue Health Monitoring System | 25 min | 1,300 copies/ml | No | 98.8% (81) | 95.6% (68) |
| DETECTR (Mammoth)<sup>47,48</sup> | E, N | NP, OP | LFIA w/ RT-LAMP and CAS-12 | None – visual | 45 min | 10,000 copies/ml | No | 95% (36) | 100% (42) |
| SHERLOCK (Sherlock Biosciences)<sup>53</sup> | E, N, ORF1ab |  | LFIA w/ RT-RPA and CAS-13a | None – visual | 70 min | 210 copies/ml | No | 100% (81) | 97% (73) |
| **Rapid Antigen Tests** |  |  |  |  |  |  |  |  |  |
| Veritor (BD) | Protein N | NS | Chromatographic immunoassay | Veritor Plus Analyzer | 15 min | 140 TCID<sub>50</sub>/swab | No | 83.9% (31) | 100% (195) |
| LumiraDx (LumiraDx) | Protein N | NS | Fluorescence immunoassay | LumiraDx Instrument | 12 min | 32 TCID<sub>50</sub>/swab | No | 97.6% (83) | 96.6% (174) |
| Sofia 2 (Quidel) | Protein N | NP, NS | LFIA | Sofia 2 Fluorescent Analyzer | 15 min | 113 TCID<sub>50</sub>/swab | No | 96.7% (30) | 100% (179) |
| BinaxNow (Abbott) | Protein N | NS | LFIA | None – visual | 15 min | 22.5 TCID<sub>50</sub>/swab | No | 97.1% (35) | 98.5% (67) |
| Biocredit (Rapigen)<sup>54</sup> | Protein N | NP | LFIA | None – visual | 8 min | N/A | No | 62% (79) | 100% (30) |
| Respi-Strip (Coris Bio)<sup>55</sup> | Protein N | NP | Dipstick LFIA | None – visual | 30 min | 5.4 pM | No | 87.9% (99) | 77.3% (132) |
| **This Work** | Protein S | OP | Aptamer-based competitive assay | Glucometer | 60 min | 4.38 pM | Yes | 100% (16) | 100% (8) |

NS = Nasal sample, NP = Nasopharyngeal swab, MT = mid-turbinate swab, OP = oropharyngeal swab

## Study Limitations

Our study has limitations. First, we selected aptamers based on literature report rather than using SELEX to develop bespoke aptamers. Even so, the aptamers chosen in this work demonstrated high affinity and specificity for SARS-CoV-2 antigens and have the advantage of prior validation and peer-reviewed reporting. Second, for testing of binding during authentic viral transmission or in clinical specimens, it should be noted that we did not attempt to lyse the virions (*i*.*e*. no detergent was added), potentially leaving a complex mixture of intact virus, partially assembled virion, and protein which may or may not be recognized due to association with neutralizing antibodies. Third, our clinical dataset was relatively small, did not contain asymptomatic SARS-CoV-2 negative individuals, and was conducted retrospectively. Larger prospective studies are needed to establish the true LOD, ROC characteristics, and correlation to clinical characteristics such as symptom status, viral load, and infectivity. Finally, as above, our assay was performed with only minimal sample preparation and only preliminary optimizations in assay conditions and workflow. Finding optimal antigen-aptamer binding conditions, including the addition of detergents, optimization of enzyme selection and reaction conditions to accommodate faster, low temperature testing, and lyophilization to increase shelf life, would each need to be addressed prior to testing at scale or approval for clinical use.

## Outlook

We developed, validated, and tested a novel SARS-CoV-2 biosensor that can sensitively and specifically detect acute viral infection from human saliva using low-cost reagents and widespread commercially available glucometers. Our preliminary results suggest such an approach could be used at scale for repeated population screening and diagnosis, however prospective clinical trials are needed to determine assay performance across a range of clinical contexts. As discussed by Paltiel and Walensky^56^, we propose that such rapid, saliva-based antigen testing can be the “*essential weapon in the fight to resume many of the activities and reopen many of the venues that comprise what we used to call normal life*.”

## Data Availability

Data is provided in supplemental.

## Acknowledgements

This work was supported in part by the National Institutes of Health (NIH) Rapid Acceleration of Diagnostics (RADx) program, a Career Award for Medical Scientists from the Burroughs Wellcome Fund to A.F.C., and a National Science Foundation (NSF) CAREER Award to D.A.H. (ECCS-1454608). We thank the UCSD Center for AIDS Research Genomics and Sequencing Core and support from the John and Mary Tu Foundation for ddPCR SARS-CoV-2 quantification. We thank Efren Sandoval, Aakash Amin, and David Becker at Helix for performing the SARS-CoV-2 RNA quantification on the clinical samples. The following reagent was deposited by the Centers for Disease Control and Prevention (CDC) and obtained through BEI Resources, NIAID, NIH: SARS-related Coronavirus 2, Isolate USA-WA1/2020, NR-52281. We thank Dr. Robert Schooley for valuable feedback and the volunteers who provided samples.

## Supplemental

**Table S1.**
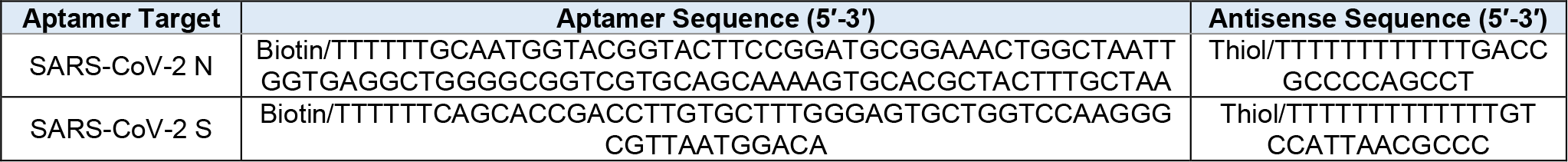
Aptamer and antisense sequences.

**Table S2.** Buffer composition used for various reactions.

| Name | Composition |
| --- | --- |
| Washing and binding buffer | 10 mM tris, 0.5 M NaCl, 1 mM EDTA, pH 7.4 |
| Sucrose buffer (4X) | 0.4 M citrate, 4 M sucrose, 20 mM MgCl<sub>2</sub>, 4 mM MgCl<sub>2</sub>, 2 mM EDTA, 11.1 mM glucose, pH 5.0 |
| Measurement buffer (2X) | 2 M sucrose, 2 mM ferrocene, 10 mM CaCl<sub>2</sub>, 2 mM MgCl<sub>2</sub>, 1mM EDTA in PBS pH 7.4 |

**Table S3.**
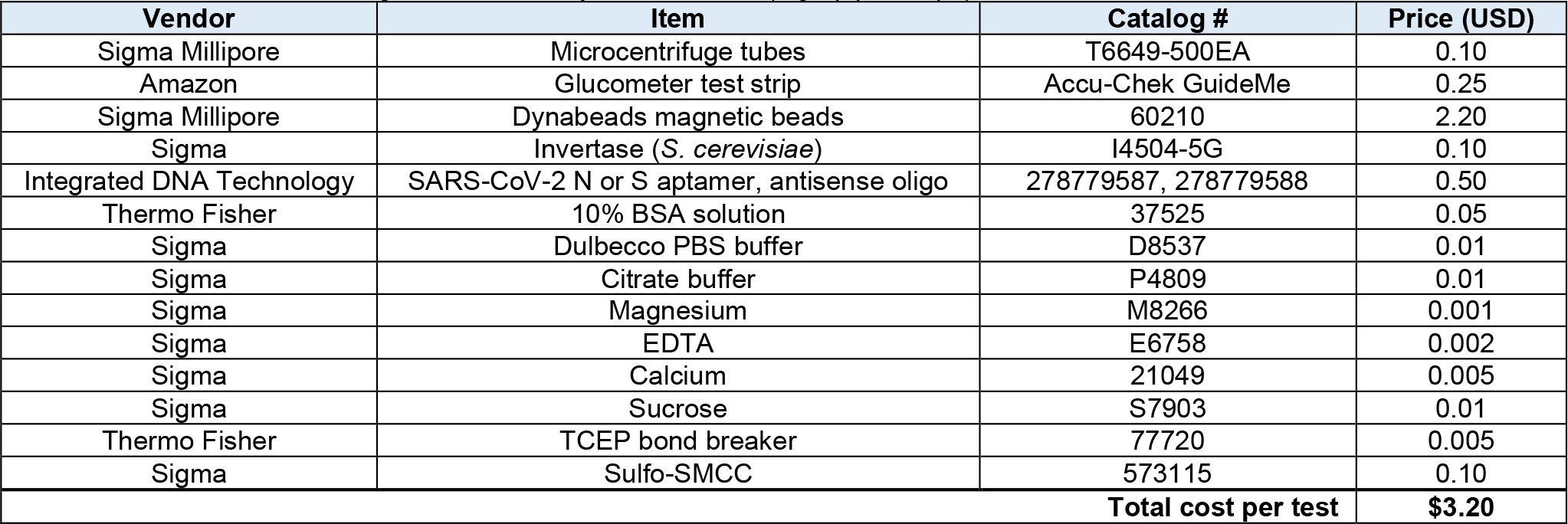
List of reagents and cost of goods per SARS-CoV-2 test. The calculated price is based on MSRP listed on vendor websites. Cost of general laboratory consumables (*e*.*g*., pipette tips) and instrumentation is not included.

| Vendor | Item | Catalog # | Price (USD) |
| --- | --- | --- | --- |
| Sigma Millipore | Microcentrifuge tubes | T6649-500EA | 0.10 |
| Amazon | Glucometer test strip | Accu-Chek GuideMe | 0.25 |
| Sigma Millipore | Dynabeads magnetic beads | 60210 | 2.20 |
| Sigma | Invertase (*S. cerevisiae*) | I4504-5G | 0.10 |
| Integrated DNA Technology | SARS-CoV-2 N or S aptamer, antisense oligo | 278779587, 278779588 | 0.50 |
| Thermo Fisher | 10% BSA solution | 37525 | 0.05 |
| Sigma | Dulbecco PBS buffer | D8537 | 0.01 |
| Sigma | Citrate buffer | P4809 | 0.01 |
| Sigma | Magnesium | M8266 | 0.001 |
| Sigma | EDTA | E6758 | 0.002 |
| Sigma | Calcium | 21049 | 0.005 |
| Sigma | Sucrose | S7903 | 0.01 |
| Thermo Fisher | TCEP bond breaker | 77720 | 0.005 |
| Sigma | Sulfo-SMCC | 573115 | 0.10 |
| **Total cost per test** | | | **\$3.20** |

**Table S4.** Demographic information, symptoms, and measurement data for cohort. All glucose values are averages from independent triplicate measurements. Symptoms and comorbidities were all self-reported. Ages rounded to nearest 10 years for confidentiality.

| ID | Age (approx.) | Sex | Days between first positive test to sample | Days between symptom onset and sample | Fever | Cough | Fatigue | Shortness of breath | Chills | Anosmia | Ageusia | Comorbidities | Hospitalized? | Ct Value | Background Glucose [mg/dl] | Δ Glucose [mg/dl] |
| --- | --- | --- | --- | --- | --- | --- | --- | --- | --- | --- | --- | --- | --- | --- | --- | --- |
| Patient 15 | 50 | M | 2 | 4 | Yes | Mild | No | No | Yes | No | Yes |  | No | 25.4 | 54.0 | 301.0 |
| Patient 18 | 30 | F | 7 | 9 | Yes | N/A | N/A | N/A | N/A | N/A | N/A |  | No | 34.7 | 91.0 | 404.0 |
| Patient 23 | 30 | F | 6 | 10 | Yes | Mild | No | No | Yes | No | No |  | No | 20.9 | 39.5 | 262.5 |
| Patient 30 | 30 | F | 4 | 3 | No | No | No | No | No | Yes | Yes | Pregnant | No | 23.1 | 42.5 | 199.5 |
| Patient 38 | 30 | M | 2 | 3 | Yes | Mild | No | No | No | Yes | Yes |  | No | 24.1 | 307.0 | 145.3 |
| Patient 40 | 30 | M | 4 | 4 | No | Mild | Mild | No | No | Yes | Yes | Asthma | No | 28.6 | 51.0 | 68.3 |
| Patient 42 | 40 | M | 5 | 7 | Yes | No | Mild | No | Yes | Yes | Yes |  | No | 22.5 | 58.0 | 217.3 |
| Patient 56 | 20 | F | 5 | 7 | No | No | No | No | No | Yes | Yes |  | No | 27.2 | 63.5 | 328.2 |
| Patient 57 | 30 | F | 4 | 6 | Yes | No | No | No | No | No | No |  | No | 26.0 | 93.0 | 359.3 |
| Patient 61 | 20 | F | 9 | 10 | No | No | No | No | No | No | No |  | No | 31.2 | 58.5 | 187.2 |
| Patient 63 | 20 | M | 7 | 15 | Yes | Mild | No | No | No | Yes | Yes |  | No | 32.2 | 181.5 | 334.2 |
| Patient 68 | 30 | F | 4 | 8 | No | Mild | Mild | Moderate | No | Yes | Yes | Asthma | No | 33.1 | 47.0 | 87.0 |
| Patient 72 | 30 | F | 3 | 8 | Yes | No | Mild | No | No | Yes | No |  | No | 21.7 | 43.0 | 122.3 |
| Patient 74 | 20 | M | 5 | 6 | Yes | No | No | No | Yes | No | No |  | No | 29.5 | 50.0 | 136.3 |
| Patient 77 | 60 | M | 6 | 5 | Yes | Mild | Mild | No | No | Yes | Yes |  | No | 23.4 | 55.0 | 201.0 |
| Patient 78 | 30 | M | 3 | 6 | No | Mild | No | No | No | No | No |  | No | 30.3 | 57.5 | 130.2 |
| Volunteer 1 | 40 | M | N/A | N/A | No | No | No | No | No | No | No |  | No | N/A | 40.0 | 27.7 |
| Volunteer 2 | 30 | M | N/A | N/A | No | No | No | No | No | No | No | Asthma | No | N/A | 35.0 | 30.0 |
| Volunteer 3 | 30 | M | N/A | N/A | No | No | No | No | No | No | No |  | No | N/A | 32.0 | 37.7 |
| Volunteer 4 | 30 | F | N/A | N/A | No | No | No | No | No | No | No |  | No | N/A | 34.0 | 17.7 |
| Volunteer 5 | 40 | M | N/A | N/A | No | No | No | No | No | No | No |  | No | N/A | 33.0 | 23.7 |
| Volunteer 6 | 40 | F | N/A | N/A | No | No | No | No | No | No | No |  | No | N/A | 39.3 | 14.0 |
| Volunteer 7 | 20 | M | N/A | N/A | No | No | No | No | No | No | No |  | No | N/A | 40.0 | 18.0 |
| Volunteer 8 | 30 | M | N/A | N/A | No | No | No | No | No | No | No |  | No | N/A | 42.0 | 25.3 |

**Table S5.**
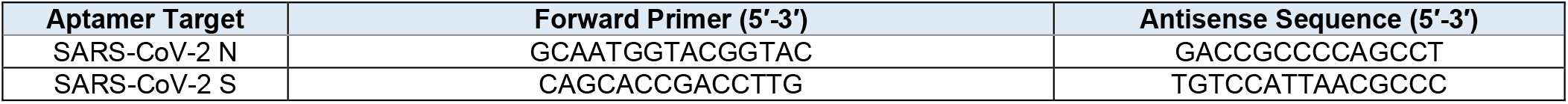
PCR primers.

**Figure S1.**
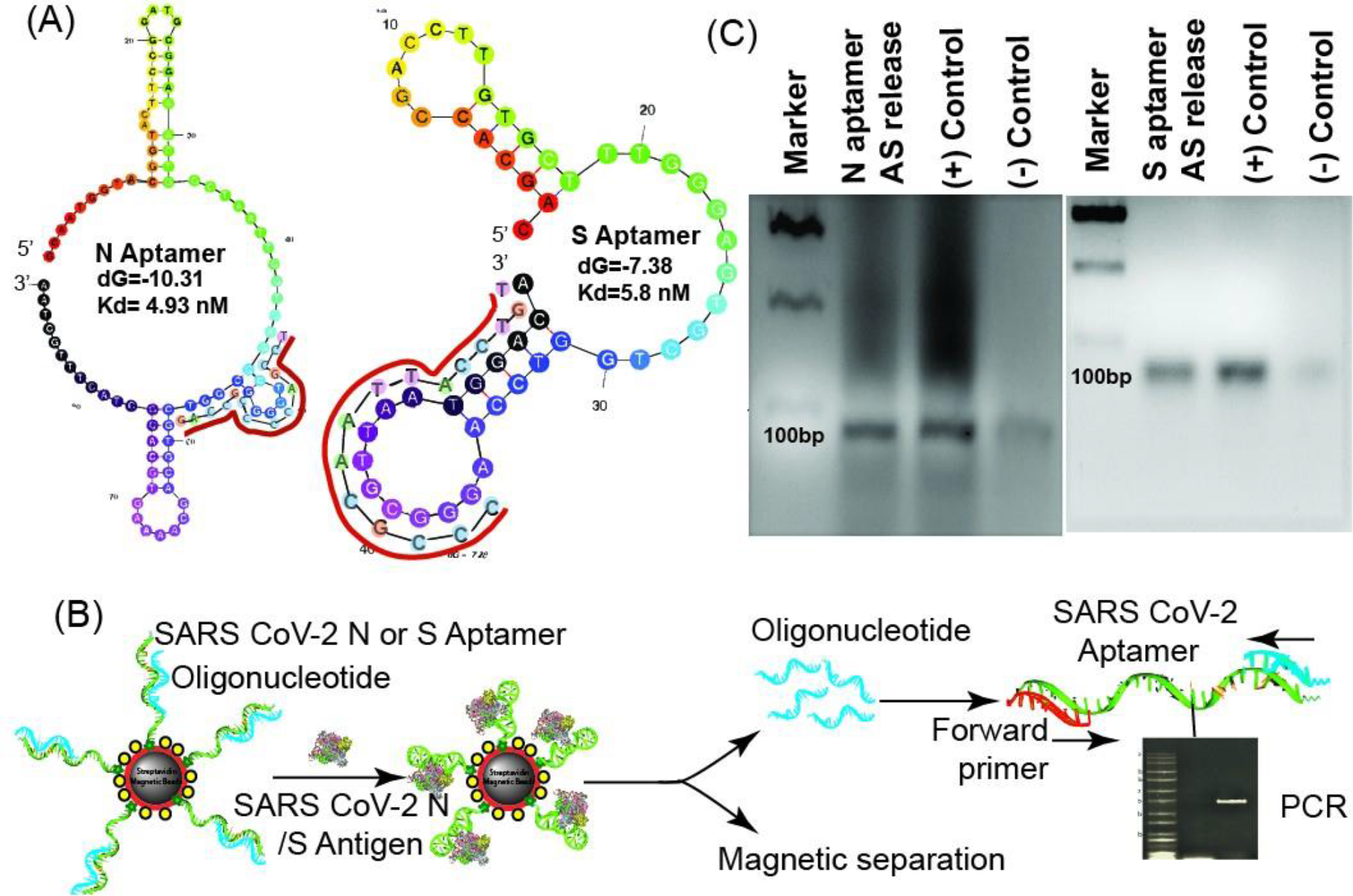
Aptamer and antisense strand displacement verification. (A) Predicted secondary structure of the N and S aptamer using M-fold. The antisense strand sequence and the binding locations are annotated in red. (B) Overview of the release study of antisense strand (blue) from aptamer (green) upon antigen binding, and validation study of the release using PCR. After magnetic separation of the MB-aptamer-antigen conjugate, the released oligonucleotide (blue) is collected from the supernatant and is added to the PCR reaction mixture with the aptamer (green) as template and the forward primer (red). PCR reaction is resolved in the agarose gel and stained with Ethidium Bromide (EtBr). (C) Antisense (AS) release study from the hybridized N and S aptamer immobilized on the magnetic beads upon antigen binding was confirmed with PCR. The PCR products were resolved in 2% agarose gel and stained with EtBr for visualization of the DNA amplicon. PCR reactions with the respective aptamer templates, forward and reverse primers was performed for (+) control. For the (-) control, only buffer (without N or S proteins) was added to the MB-aptamer-antisense (AS) complex. A 100 base-pair ladder was also resolved as molecular-weight marker.

**Figure S2.**
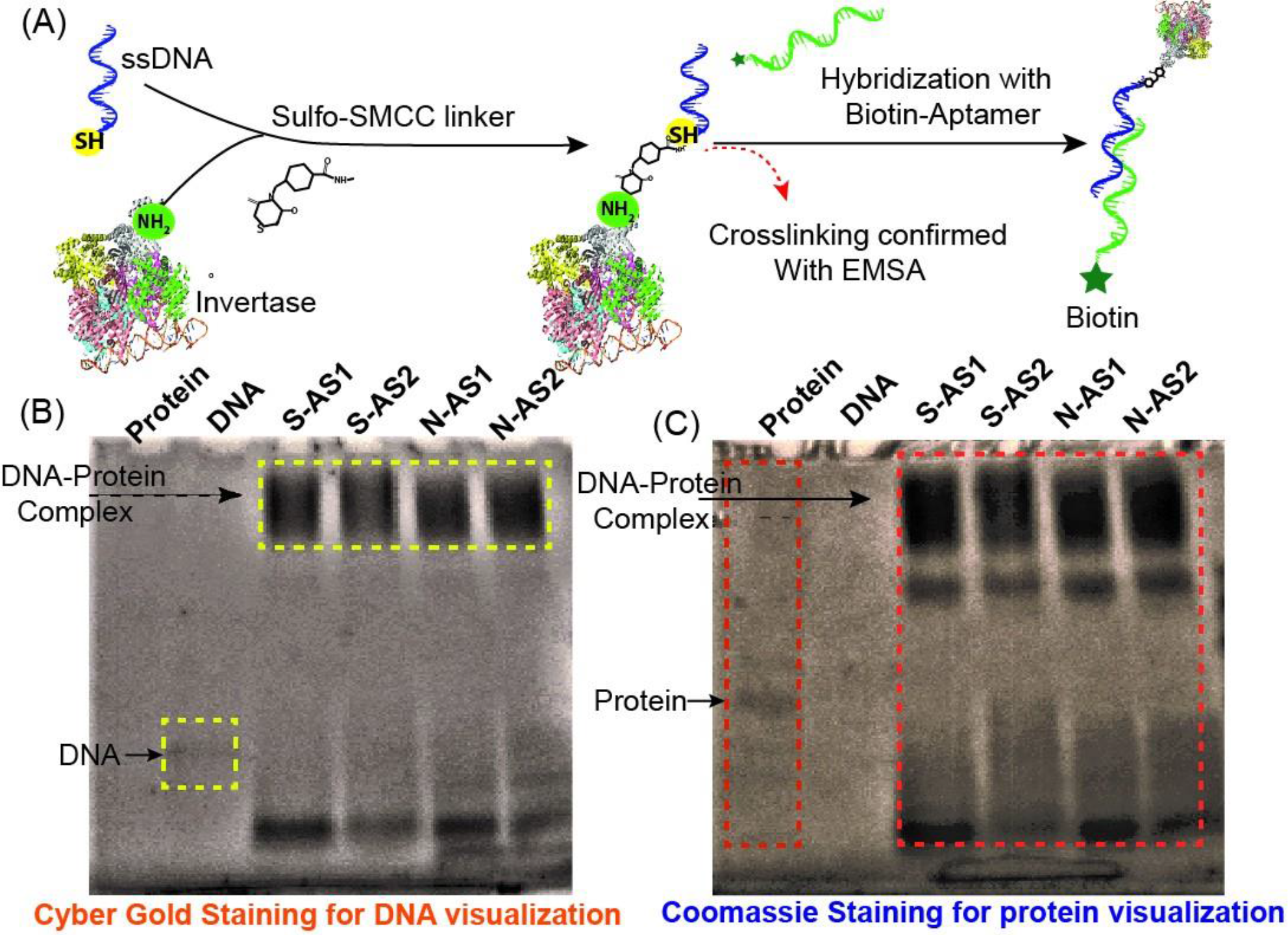
Conjugation of antisense-invertase enzyme. (A) Overview of cross-linking antisense oligonucleotide with invertase and hybridization with the respective biotinylated-aptamer. (B) N-antisense (N-AS) and S-antisense (S-AS) conjugation with invertase was verified by an electrophoretic mobility shift assay (EMSA). The conjugates were resolved in 4-20% gradient native acrylamide gel for 2 hours in 1X TBE buffer at 100 V. Unconjugated DNA and the invertase protein were run as controls. S-AS1, S-AS2 and N-AS1, N-AS2 depict two different concentrations of antisense-invertase enzyme conjugates. The gel was stained with (B) Cyber Gold, followed by (C) Comassie brilliant blue for DNA and protein staining, respectively. Higher migrating bands were detected at the same spot with both the DNA and protein specific dyes, thus indicating successful conjugation.

**Figure S3.**
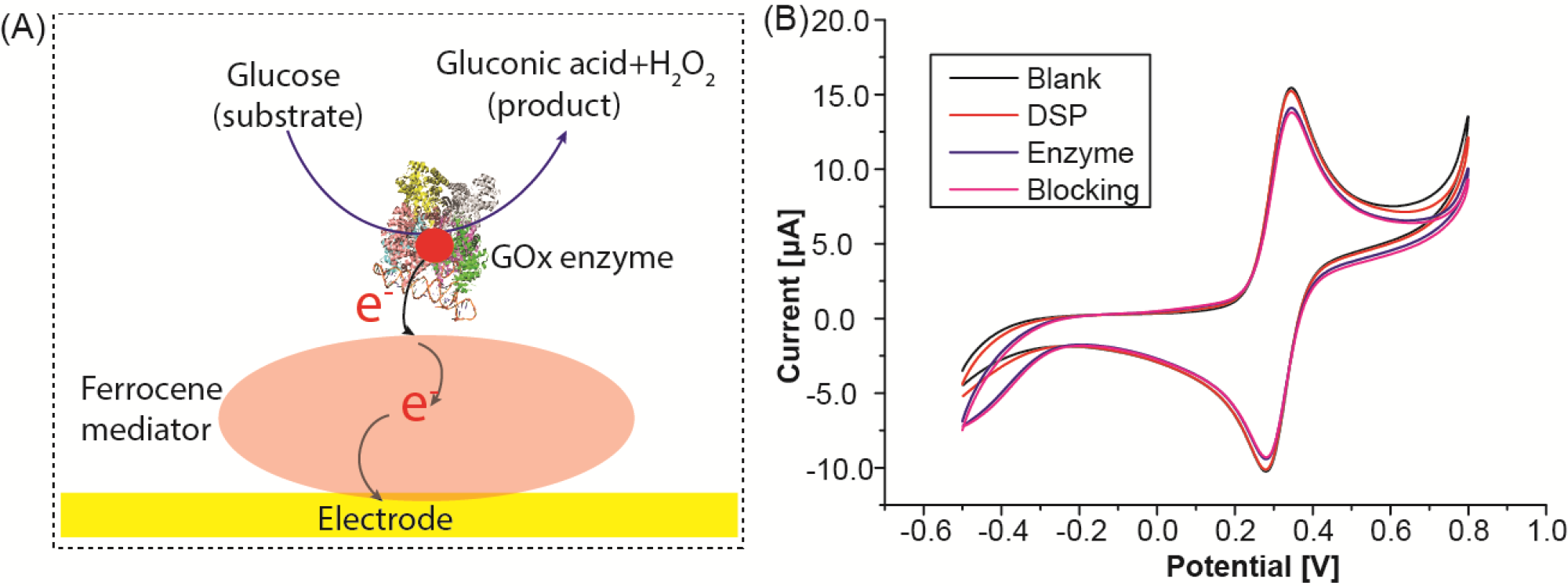
Custom glucose sensor fabrication and assessment. (A) Overview of glucose sensor operating principle. (B) Characterization of the sensor fabrication showing a gradual reduction in current after layer by layer immobilization of DSP, enzyme (GOx), and blocking confirming successful stepwise assembly.

**Figure S4.**
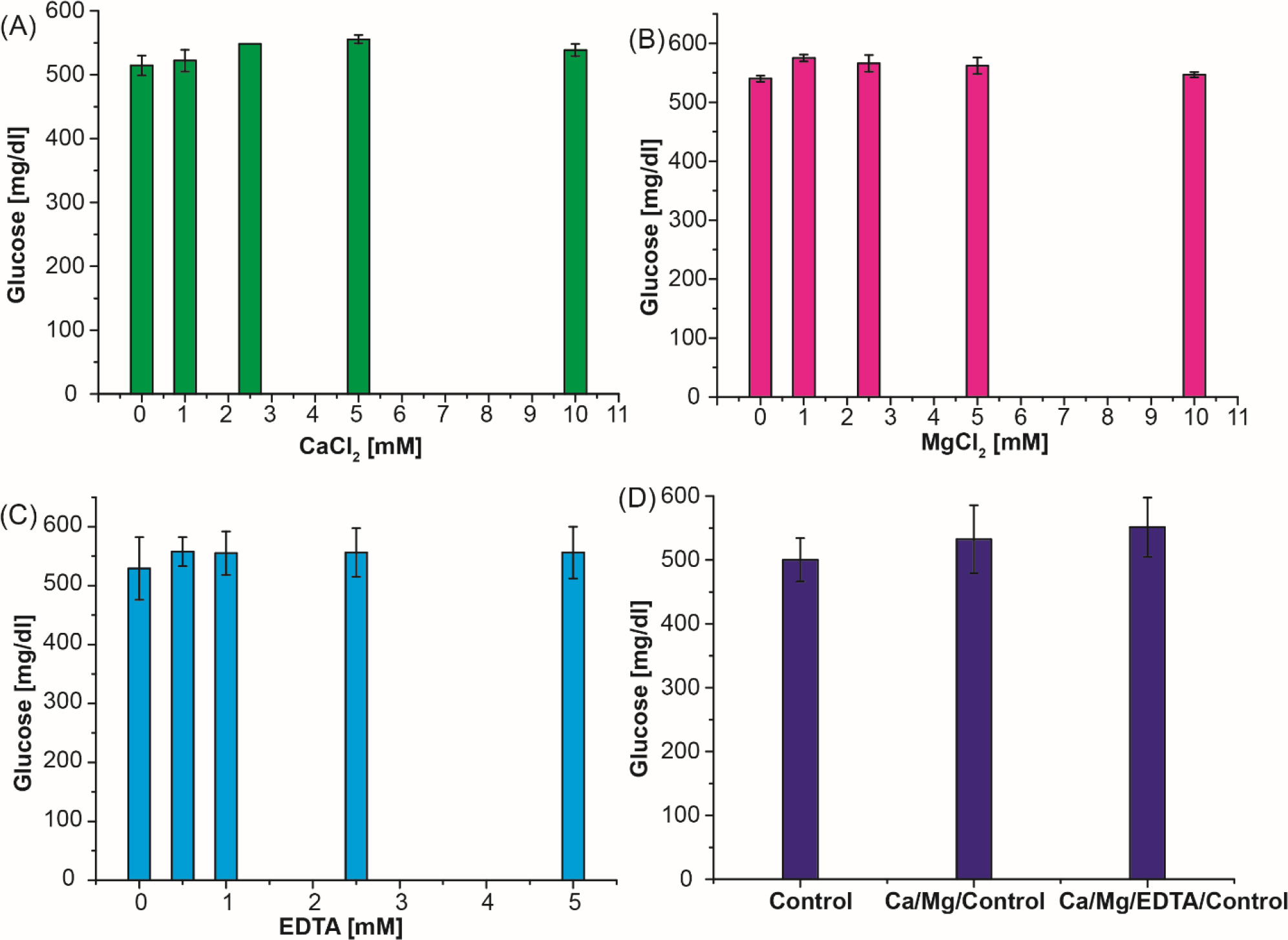
Ion and salt optimization for amplification buffer. Metal ions and salts have a crucial role on enzyme activity. Optimization study with concentration of different salts (A) Ca and (B) Mg, and (C) EDTA on enzyme activity. The effect of EDTA on metal ion chelation was observed to increase invertase activity without any inhibitory effect. (D) Optimization using best conditions from (A-C). These optimization studies were performed using 1.0 M sucrose with 1 µM invertase enzyme at RT for 30 min. Maximum invertase activity was observed with 5 mM CaCl_2_, 1 mM MgCl_2_, and 0.5 mM EDTA.

**Figure S5.**
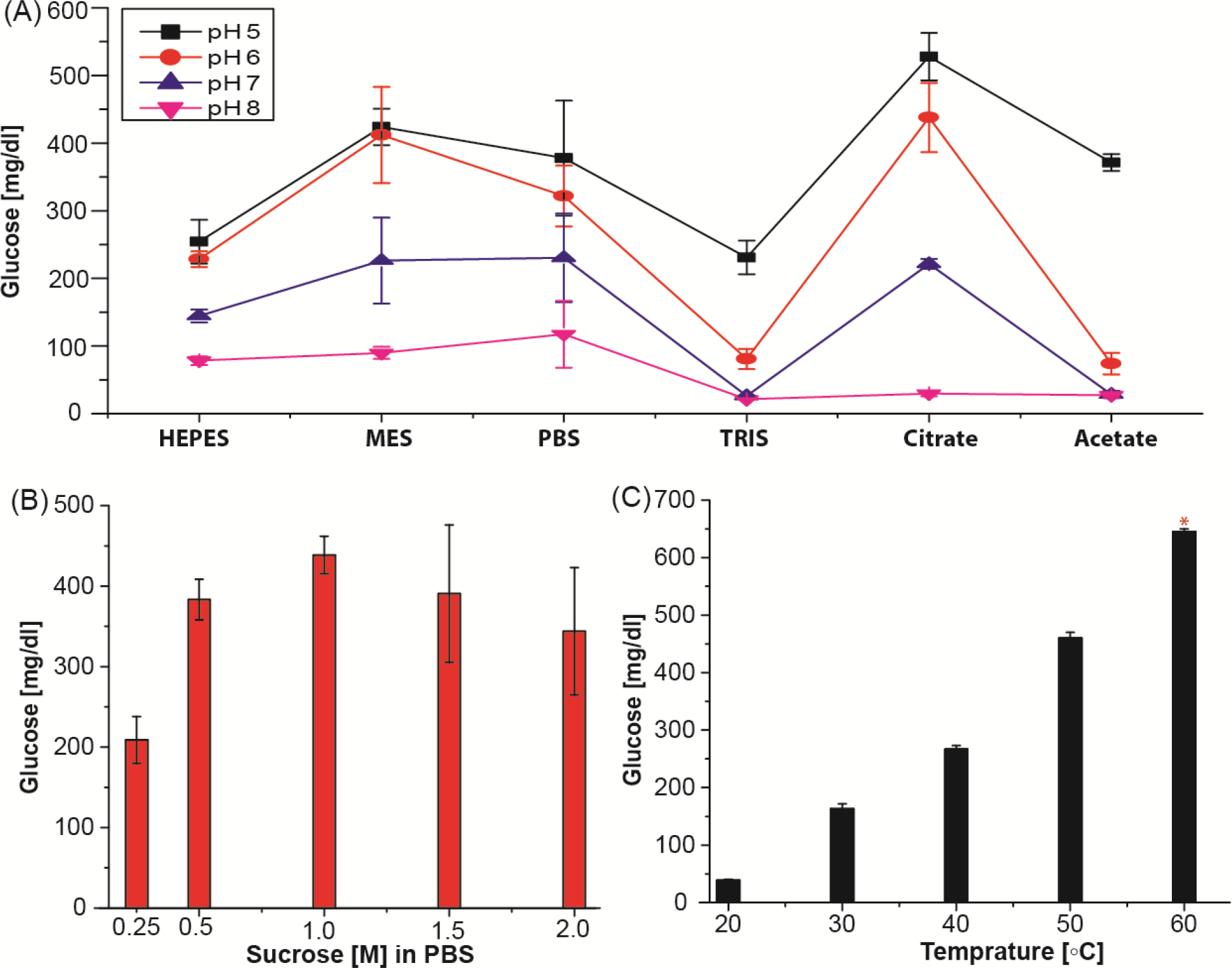
Amplification buffer optimization. (A) Buffer optimization with 0.1 M buffers at different pHs. (B) Substrate concentration optimization in 1× PBS and (C) temperature study in 0.1 M citrate buffer pH 5.0. * indicates that the sample was diluted 2-fold due to the limited dynamic range of the glucometer. All the enzyme optimization studies were performed using 1.0 M sucrose with 1 µM invertase enzyme at RT for 30 min.

**Figure S6.**
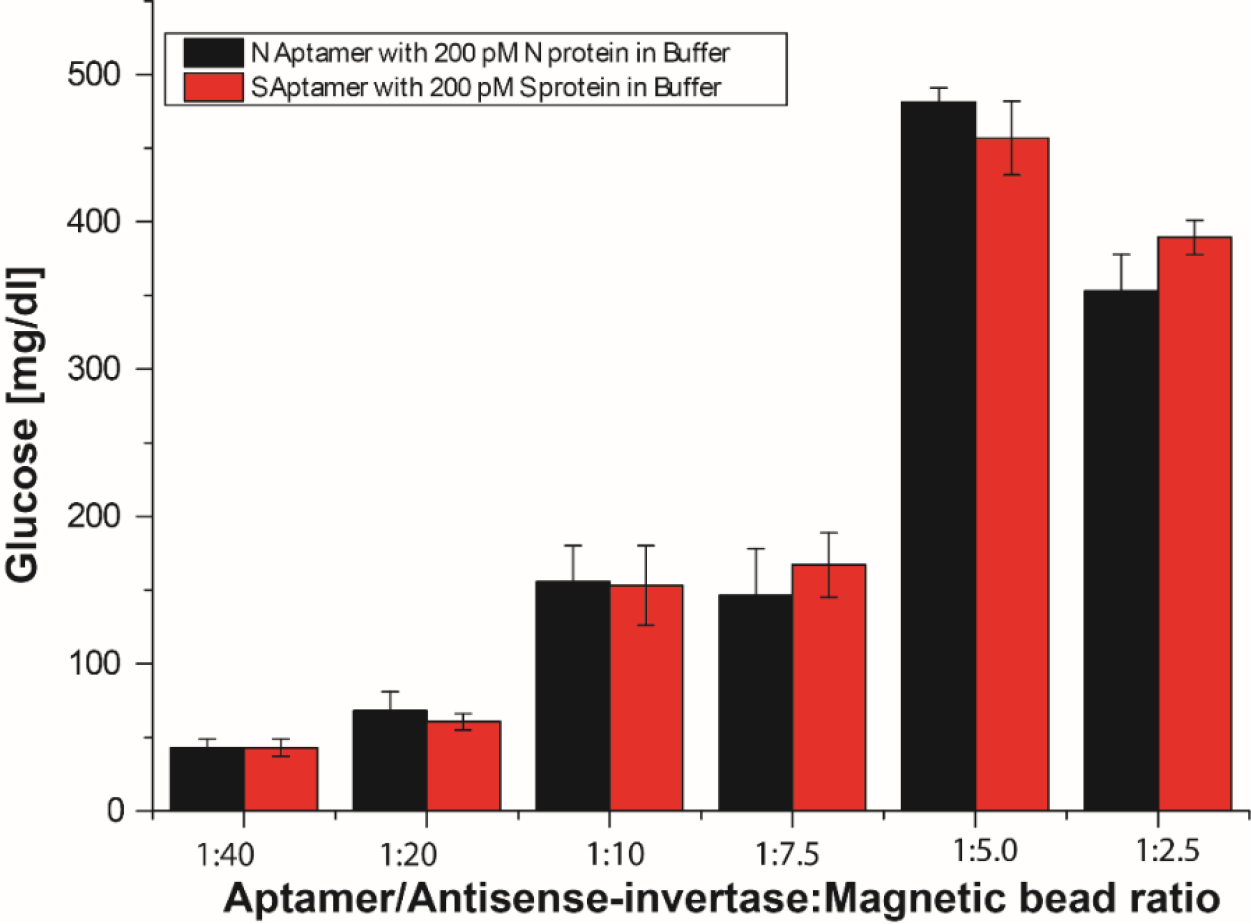
Effect of aptamer-antisense-invertase conjugate to streptavidin coated magnetic bead ratio on antigen binding. The efficiency of the assay system was varied from 1:40 to 1:2.5 and saturated at 1:5 (60 µg of aptamer: 300 µg of MBs) for both aptamers.

**Figure S7.**
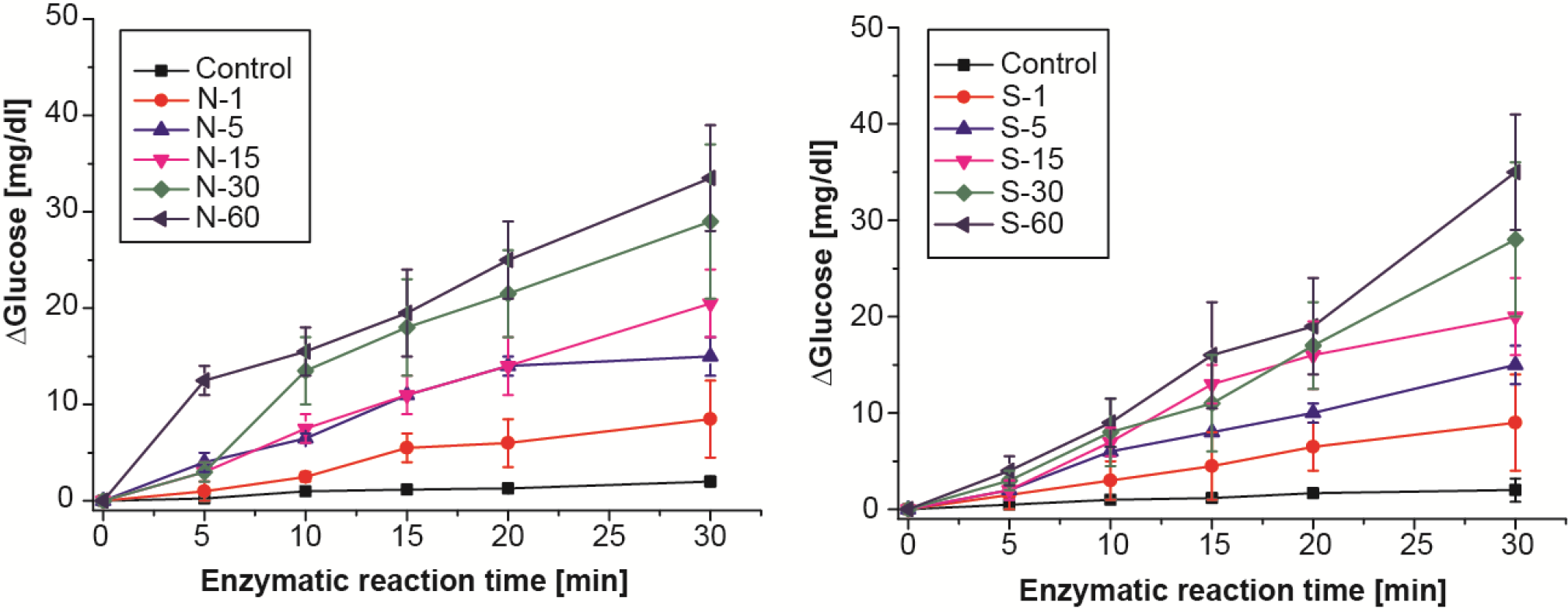
Enzymatic reaction kinetics. Aptamer binding and enzymatic reaction time optimization for (A) N and (B) S aptamer complex systems in buffer. N or S aptamer complex was incubated against respective target for 1 to 60 min and corresponding invertase activity assay performed up to 30 min at 5 min intervals. The control experiments were performed similarly in the absence of antigen. The assay was performed using the optimal ratio of aptamer/antisense-invertase: MB (1:5) with 300 μg of N or S aptamer/antisense-invertase-MB complex. As expected, the longer the incubation time for aptamer-target interaction, substitute the higher concentration of antisense-invertase conjugate and enhance the rate of sucrose conversion.

**Figure S8.**
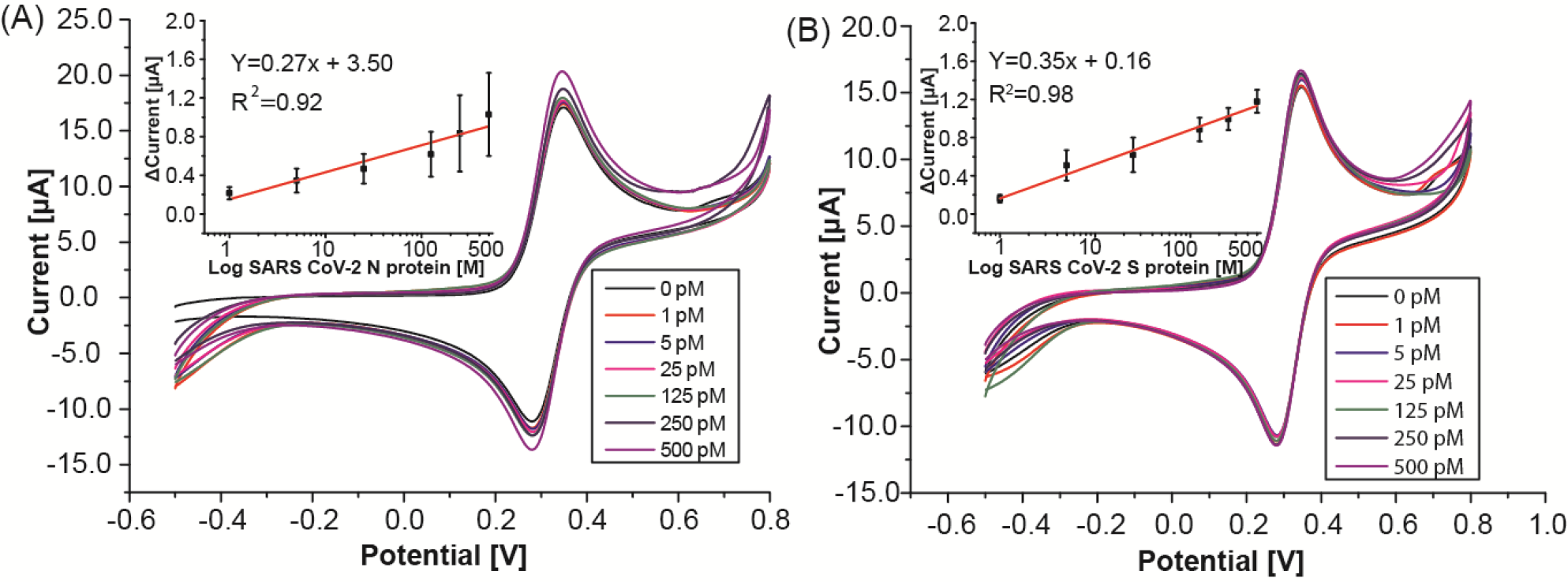
Measurement of SARS CoV-2 antigen with custom glucose sensor. Measurement results from (A) N and (B) S SARS-CoV-2 protein spiked in buffer at various concentrations (1-500 pM). Inset shows calibration plot after background subtraction. Measurements were performed in 1x measurement buffer with ferrocene mediator to facilitate the electron transfer from enzyme redox center to electrode surface. An incremental shift in the oxidation peak of the voltammograms at higher concentrations. The calculated LODs are 0.71 pM and 0.34 pM for SARS CoV-2 N and S protein, respectively.

**Figure S9.**
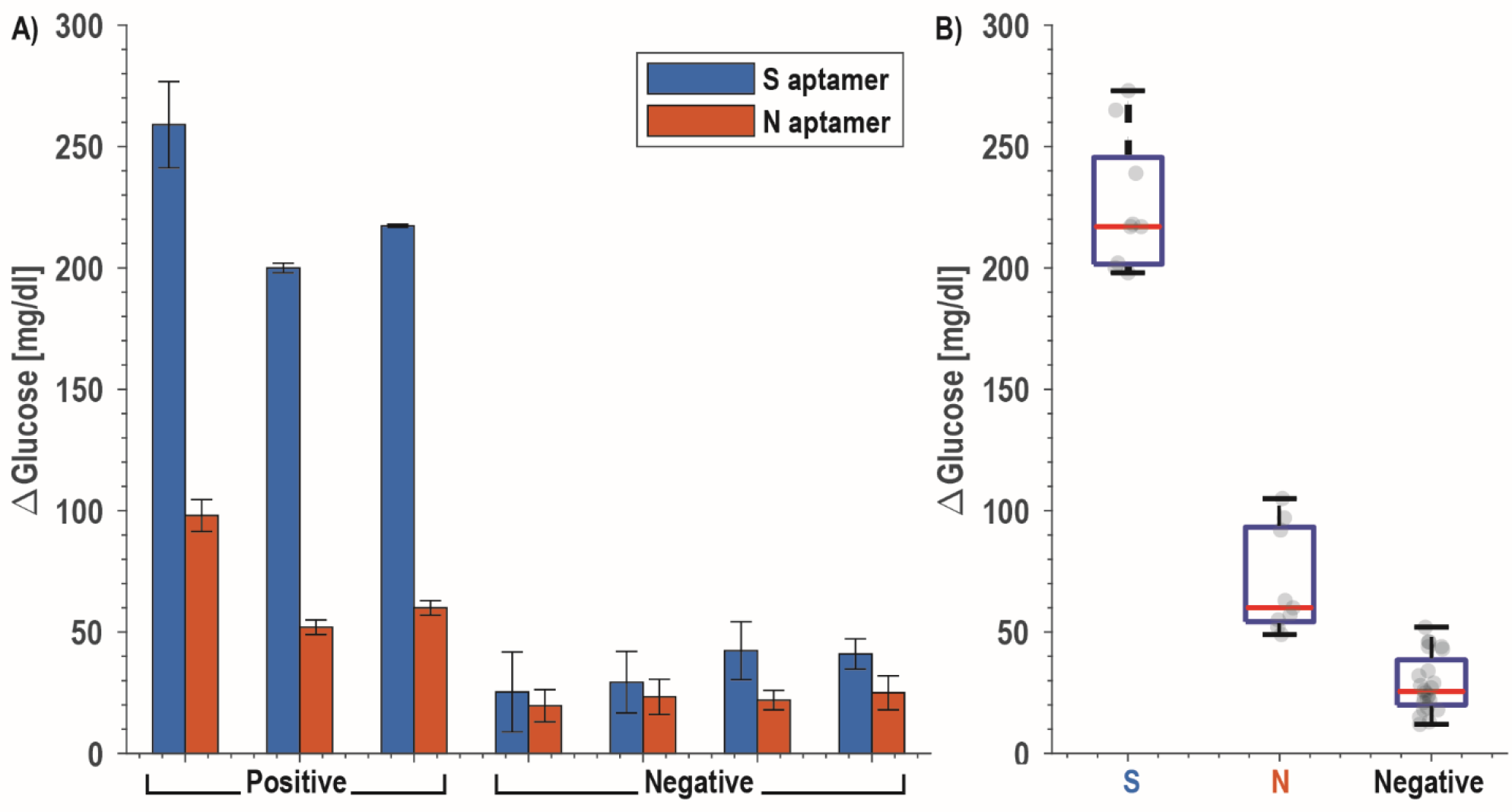
CoVID-19 clinical saliva samples. (A) Measured data from confirmed positive patients (*N*=3; patients 23, 30, and 42) and healthy volunteers (*N*=4) for paired N and S aptamers. Detection of SARS CoV-2 N protein was performed with the addition of 1% Triton to ensure the release of the nucleocapsid protein. (B) Box and whisker plot.

**Figure S10.**
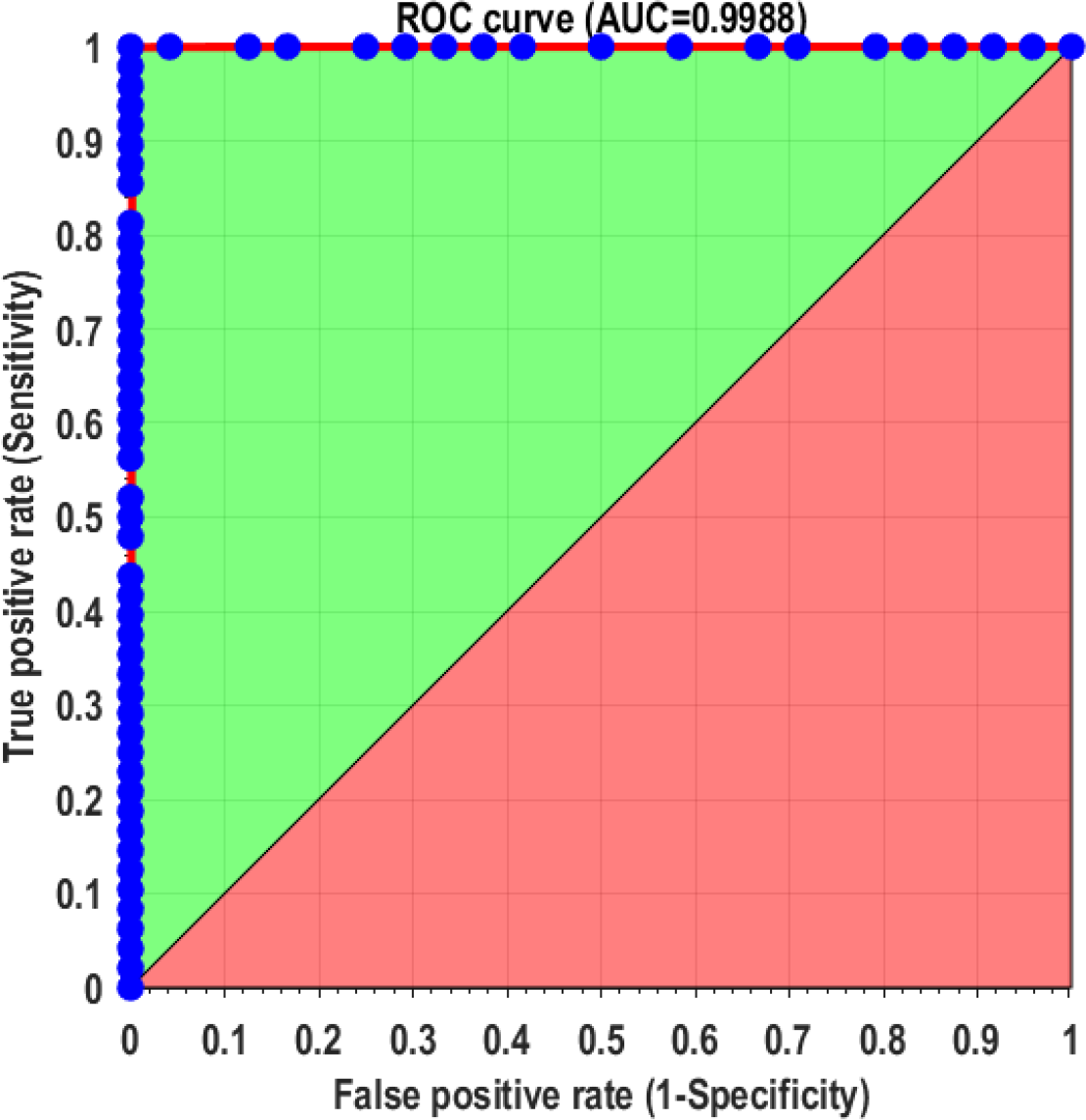
Receiver operator characteristic (ROC) curve. All individual measurements were plotted using MATLAB and function provided by Giuseppe Cardillo (http://www.mathworks.com/matlabcentral/fileexchange/19950).

